# Can non-routine data collection help decision-making for *gambiense* sleeping sickness? Using active adaptive management to assess the potential of vector control

**DOI:** 10.64898/2026.07.27.26359007

**Authors:** Rob Sunnucks, Inaki Tirados, Erick Mwamba Miaka, Emma L Davis, Kat S Rock

**Affiliations:** Mathematics Institute, The University of Warwick, Academic Loop Road, Coventry, CV4 7AL, U.K. EPSRC & MRC Centre for Doctoral Training in Mathematics for Real-World Systems, University of Warwick, Coventry, United Kingdom; Zeeman Institute for Systems Biology and Infectious Disease Epidemiology Research, The University of Warwick, Academic Loop Road, Coventry, CV4 7AL, U.K; Department of Vector Biology, Liverpool School of Tropical Medicine, Liverpool, U.K; Programme National de Lutte contre la Trypanosomiase Humaine Africaine (PNLTHA), Kinshasa, D.R.C; The Department of Statistics, The University of Warwick, Academic Loop Road, Coventry, CV4 7AL, U.K

## Abstract

*Gambiense* human African trypanosomiasis (gHAT) is a vector-borne disease, spread by tsetse, found in West and Central Africa. It has been targeted for elimination of transmission (EoT) by the World Health Organization: the reduction to zero of the incidence of infection, with minimal risk of reintroduction. Many high-risk regions have implemented vector control (VC), in addition to medical interventions, to reduce the spread of gHAT and reach EoT. There exists uncertainty around the effectiveness of VC interventions, and so the impact of VC can be measured through deploying traps, which estimate changes in the tsetse population. In this study, we use mathematical transmission modelling to assess the added monetary value of entomological monitoring data in strategy selection for two health zones of the Democratic Republic of Congo, illustratively chosen because of their different levels of uncertainty on VC effectiveness.

We used active adaptive management (AAM)—the process of updating our strategy based on collected data (i.e. choosing to continue or stop VC based on perceived effectiveness), where posterior distributions on VC effectiveness were calculated with an adaptive MCMC method. Next, we used an established gHAT transmission model along with the policy objective of maximising the intervention’s net monetary benefit to calculate the value gained by various monitoring strategies.

We found that the strategy that included VC and twice-yearly entomological monitoring was cost-effective at a willingness to pay threshold of $153 for the health zone of Mulumba, where we were able to determine the optimal monitoring strategy, but it was not cost-effective in the health zone of Vanga.

This study is the first application of AAM to monitoring data for a vector-bone infection, presenting a novel framework for quantitative assessment of how VC monitoring could be used to improve the cost-effectiveness of interventions in other regions.

**Author summary:** *Gambiense* human African trypanosomiasis (gHAT) is a disease in West and Central Africa spread by tsetse. A key method to combat the spread of the disease is the deployment of tiny targets which kill tsetse. Monitoring of the tsetse population can allow us to ascertain the effectiveness of tiny targets in a given area and use this information to inform whether deployments should continue. However, the usefulness of this monitoring has been called into question. Here, we determine the monetary benefit of this monitoring in reducing the burden of gHAT and lowering costs.

We calculated the benefit of conducting twice-yearly monitoring over a two-year period of vector control, and how this changed for different locations and different monitoring strategies. We found these efforts were cost-effective at a willingness to pay threshold of $153 for the health zone of Mulumba, where we were able to determine the optimal monitoring strategy, but it was not cost-effective in the health zone of Vanga.

This work will provide a framework to ascertain the benefit of further monitoring efforts in more complex decision-making concerning gHAT, and could also be applied to other infectious diseases.

## Introduction

*Gambiense* human African trypanosomiasis (gHAT), is on the World Health Organization’s (WHO) list of neglected tropical diseases (NTDs) and is endemic in 24 countries of West and Central Africa. In 2024, it made up more than 94% of reported cases of sleeping sickness (with the remaining cases attributed to *rhodesiense* HAT) [1]. During the first (haemolymphatic) stage, the disease manifests with non-specific signs and symptoms, such as joint pain, fever, and headaches, before Trypanosoma parasites cross the blood–brain barrier and affect the central nervous system. Once the central nervous system is affected (second, or meningoencephalitic, stage), the clinical presentation shifts towards neurological and psychiatric manifestations, including sleep disturbances, behavioural and personality changes, tremors, and dysarthria, with untreated cases often progressing to coma and death [2]. Individuals with gHAT can be infected for years without gHAT-specific signs or symptoms. When evident symptoms emerge such that they are recognisable as gHAT, the disease is often advanced, with the central nervous system already affected [3].

gHAT is a vector-borne disease, caused by a protozoan parasite *Trypanosoma brucei gambiense* and spread by species of the genus Glossina (Diptera), commonly known as tsetse [3]. These blood-feeding flies have a unique reproductive biology. Unlike most insects, tsetse reproduce through adenotrophic viviparity: eggs hatch within the female’s body, and the developing larva is nourished throughout development by secretions from specialised milk glands. Larval development is completed within the mother’s uterus, after which a single fully developed third-instar larva is deposited approximately every 7–10 days. The larva immediately pupates in the soil, where the puparium remains for approximately 30–40 days, depending on temperature, before the adult fly emerges [4]. As a consequence of this reproductive cycle, tsetse have a low reproduction rate but have a high probability of surviving to adulthood [4]. Therefore, tsetse are known as *k* strategists, which are species with stable populations that tend to a natural carrying capacity of their environment [5].

Efforts to combat the spread of gHAT have been in place for many years since the early 20th century, however, the effects of decolonisation and concerns for the environment due to the use of substances such as DDT resulted in a lack of implemented interventions against gHAT, which is attributed to the resurgence in cases of sleeping sickness in the latter half of the 20th century [6]. The more recent efforts to combat gHAT began in the late 1990s and have primarily taken the form of medical interventions, including mass screening of the population [2]. Currently, before treatment is administered, a confirmation test must be done in addition to the initial screening tests (card-agglutination trypanosomiasis test / rapid diagnostic tests), as these initial tests have a low positive predictive value [7], due to the very low prevalence of gHAT. Confirmation must occur because the drugs are not considered sufficiently safe to administer without certainty of true infection, and because of the high proportion of false positives. These interventions led to the reported annual worldwide number of new cases of the chronic form of gHAT falling by 98%, from 27,862 to 546 between the years 1999 and 2024 [1]. Passive screening consists of reporting from and treatment given in health facilities when people seek out a diagnosis themselves. Active screening consists of teams who go out into the health zone and test the at-risk population for gHAT, then provide treatment to those who are confirmed to be positive. Both passive and active screening for gHAT have occurred in the DRC.

The WHO has targeted gHAT for the elimination of transmission (EoT) by 2030, which is defined as the reduction to zero of the incidence of infection in a defined geographical area, with minimal risk of reintroduction, as a result of deliberate efforts [8]. The WHO has two global gHAT EoT indicators in order to determine whether this has been reached. These are having zero HAT cases reported, and 15 countries being verified for interruption of transmission [8]. The country-level criteria is having zero g-HAT human cases detected in all of the country’s health districts for a minimum of 5 consecutive years, along with indicators of effective surveillance [9]. Most cases of gHAT are located in the Democratic Republic of Congo (DRC), with 60.4% of reported gHAT cases in 2024 occurring in the DRC [1] and, as such, it is a key country in the global elimination of gHAT. Efforts in DRC for the last 25 years were originally comprised of different modes of screening and treatment for gHAT, but since 2015 vector control (VC) against tsetse has also been used in select health zones, in former Bandundu province (chosen for the high gHAT prevalence [10]) to further reduce gHAT transmission, and hence future cases, by reducing the tsetse population which carries the parasite between hosts [11].

Several methods of VC exist to control gHAT, including stationary baits, and sterile insect technique [12]. The low livestock (specifically bovine) presence in the DRC [13] makes approaches based on insecticide-treated cattle not viable, even if such methods may be cost-effective in other regions [12]. We therefore focus on stationary baits, specifically the deployment of Tiny Targets, as they are lower cost per area protected than other viable methods for the DRC [14]. Tiny Targets are small (0.125 m^2^) panels of blue polyester with a secondary panel of black polyethylene netting containing the insecticide deltamethrin. Tsetse are attracted to the specific colour of blue in these targets and, as they approach, they collide with the netting, picking up a lethal dose of the insecticide [11]. Although no insecticide resistance to deltamethrin has been reported in tsetse, the efficacy of the insecticide declines after approximately six months under field conditions [15]. This, combined with the impossibility of treating the entire population (unless the intervention is conducted in an isolated area), makes population recovery following a single target deployment likely. Therefore, repeated VC interventions are likely to be required to keep the tsetse population suppressed, with tiny target deployments in the DRC occurring every six months, to account for the decline in the performance of the insecticide after this period [11].

Previous Tiny Target deployments in different settings have been effective, achieving tsetse density reductions of between 60 and 100% after one year of deployment [11, 16–19]. However, these interventions were generally conducted in carefully selected active foci where conditions were considered favourable for successful tsetse suppression. Consequently, their reported effectiveness may not be directly extrapolable to large-scale interventions across the geographically, ecologically, and epidemiologically heterogeneous landscapes of the DRC, where lower or more variable levels of effectiveness can reasonably be expected [11].

To ascertain the success of an implementation such as VC, either an implementation indicator, such as the impact of VC on tsetse density, must be measured, or the human case data itself can be used. However, for gHAT, there is a long delay, typically over a year [20, 21], between an individual becoming infected and the reporting of that case, which can impede rapid determination of the effectiveness of an intervention such as VC. To measure the impact of Tiny Target deployments on tsetse density, pyramidal traps can be deployed and then revisited at 1–2 day intervals to capture tsetse and estimate their density [22]. The reduction in fly density from catches at multiple time points then serves as a proxy for the effectiveness of the VC.

Modelling can be used to help predict how the transmission of gHAT infection will change over time, allowing comparison of different intervention strategies and their ability to achieve EoT [23]. Combined with health economics, this can be used to ascertain the costs and cost-effectiveness of different strategies across different regions. But, greater uncertainty regarding future gHAT transmission and the effectiveness of available interventions reduces policymakers’ confidence in selecting optimal control strategies. Therefore, the question is whether the implementation indicator of tsetse catch data, which has never before been used directly for policy decision making, could be used to reduce our uncertainty and improve strategy selection For example, if we are uncertain about the effectiveness of VC in a new region where it is never been deployed before, we could first plan to do VC for a period of time and use data from tsetse traps to monitor the apparent reduction in density. Following the initial implementation and monitoring period, the decision to continue or discontinue the vector control intervention can be based on its measured effectiveness. Mathematical transmission models can take the monitoring strategy into account before the data is collected to determine if it is a good use of resources to put monitoring in place in this way; this is known as active adaptive management (AAM). To date, AAM has been used in some epidemiological analyses, particularly for livestock infections [24–26], however, the AAM methodology has not yet been explored for gHAT or any other NTDs, nor has it been used to consider varying the type of monitoring strategy (e.g. collecting more or less data).

In this study, we aim to determine the value gained from tsetse monitoring for gHAT decision making through an extension to the AAM framework. This required exploring the interface between epidemiological modelling, Bayesian inference and health economics to calculate the costs and benefits of undertaking AAM. To provide concrete examples for this methodology, we focused on two health zones (administrative regions of around 150,000 people) in the DRC, which, as of 2024, have had no previous VC, but have been highlighted in a previous modelling study as being unlikely to meet the EoT goal by 2030 without additional interventions [27]. Mulumba health zone in Lomami province had 78 reported cases in 2001 and 2 in 2020; none of the health zones in this region have previously had VC and so there is limited knowledge about how effective it might be in this geographical context. In contrast, Vanga health zone has a higher endemicity, with 73 reported cases in 2001 and 3 in 2020, but is near health zones which have had VC in recent years, including one with 90% reduction after one year [11]. In each health zone, we considered how differing tsetse monitoring strategies and our initial uncertainty about the impact of VC can affect the value of trap data for decision making and ascertain the optimal number of traps to use during monitoring visits.

This work is novel in expanding the AAM framework to include more complex monitoring data, building off the existing work done on AAM, as well as on work done to model the transmission of gHAT [27]. In previous AAM studies, the authors considered learning additional information from binomially distributed outcome data (such as treatment or vaccine effectiveness) and were able to use this to update their prior beliefs about the intervention. Since the priors had a beta distribution, they could be neatly updated with the monitoring data due to the conjugate nature of the beta and binomial distributions [24, 26]. In this study, the data is more complex, in that multiple parameters must be fit, and we lack a conjugate prior to our distribution of data. As such, we utilise a Markov Chain Monte Carlo (MCMC) algorithm to sample from the posterior distribution instead. Additionally, this work also considered additional costs and optimisation of the non-routine, monitoring data collection; previous publications did not factor costs into monitoring strategies.

We showed that, in Mulumba, which has more prior uncertainty in probable effectiveness of VC, for some cost-effectiveness thresholds, it may be valuable to use monitoring data to quickly gain information about the impact of the new intervention and therefore assess whether to continue or stop after two years. We also determined the optimal number of traps to use during monitoring visits in Mulumba. Additionally, we showed that in Vanga, where there is already a reasonably strong belief about the probable effectiveness of VC, the monitoring data does not provide enough additional information for our decision-making to alone justify the cost of the monitoring. This work provides a foundation for applying the AAM modelling approach to other health zones and intervention combinations, while offering guidance on its adaptation to other types of complex data and infectious diseases.

## Materials and Methods

### Example gHAT setting

In this analysis, we selected Vanga health zone in Kwilu province, in the Bandundu Nord coordination (coordinations are geographical areas based on the location of gHAT logistical units), and Mulumba health zone in the Kasaï Orientale coordination of the DRC as exemplar locations to demonstrate our AAM methodology (see Figure 1). Bandundu is a hilly area, with mix of secondary forest and crops. Because that difference in altitude, there are several small streams, which may also be flanked by riverine forest. Kasai is generally flatter than Bandundu, with much less vegetation and small rivers and so less suitable for tsetse. In both of these health zones, EoT is currently unlikely to occur by 2030 under their current medical-only strategies, with probabilities of EoT by 2030 being placed at 26% for Vanga and 15% for Mulumba [27], and neither health zone has had previous VC. Vanga had an estimated population of 335,422 in 2020, and in total, there are 93 km of large rivers (those with an average long-term discharge estimate for river reach of 20*m*^3^*s*^−1^) along which large-scale VC deployment has been deemed suitable [27]. Since 2000, Vanga has had regular but variable active screening to detect cases, with a maximum coverage of 21% in 2016. Mulumba had an estimated population of 337,594 in 2020, and in total there are 118 km of large rivers along which VC can be conducted [27]. Since 2000, Mulumba has had variable active screening to find and detect cases, with a maximum coverage of 6%.

**Fig 1.**
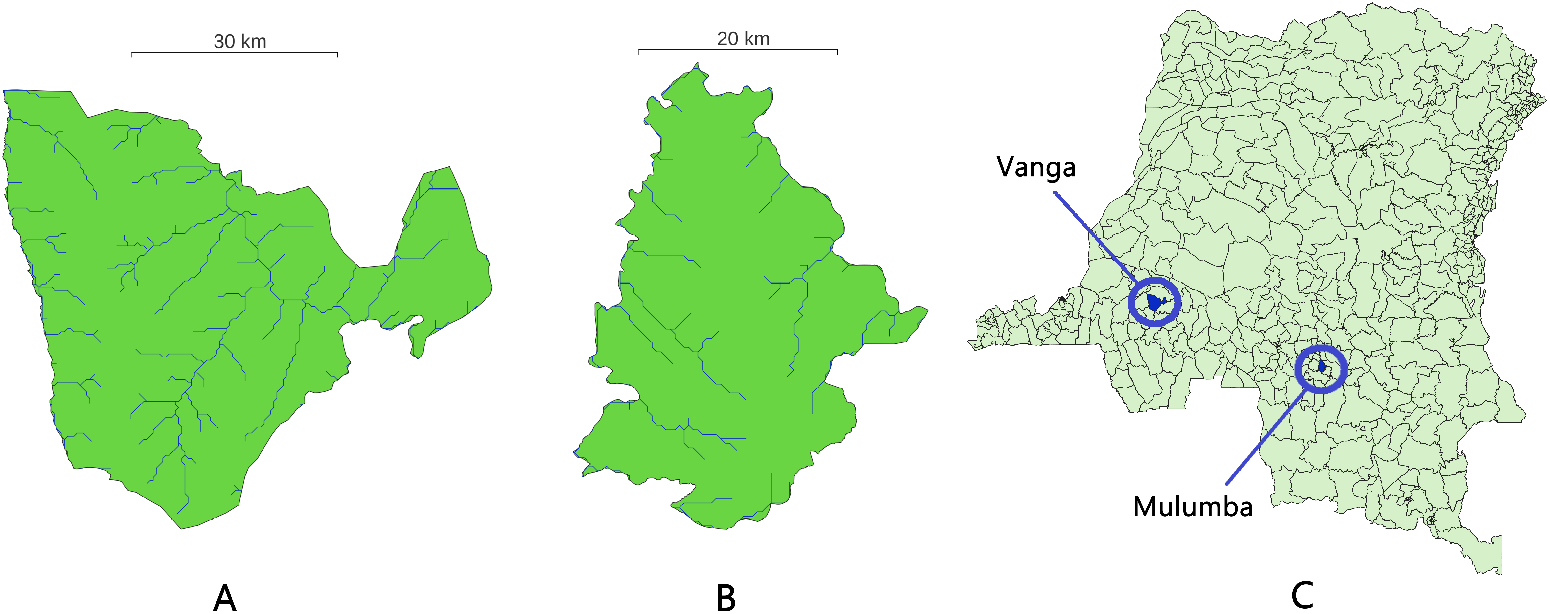
Maps of the two exemplar health zones in the DRC that we consider for this demonstration of the AAM approach to use tsetse catch data as part of strategy decision making for gHAT. A (Vanga) and B (Mulumba) are shown separately with blue lines indicating rivers. C shows the position of these health zones with the DRC. Shapefiles used to produce these maps are available under a CC-BY 4.0 licence (https://data.grid3.org/datasets/GRID3::grid3-cod-health-zones-v8-0/about) and river information is available under a CC-BY 4.0 licence from HydroATLAS (https://www.hydrosheds.org/hydroatlas).

11 health zones in Bandundu Nord and Sud coordinations have had previous VC interventions [11]. This means future interventions across the former Bandundu province may have similar success in tsetse reductions in other health zones in these coordinations, due to their similarity. The Yasa Bonga health zone, in Bandundu Sud coordination, adjacent to Bandundu Nord coordination, has had previous Tiny Target deployments with both costs and monitoring data being well documented [11, 28]. We expect the effectiveness of an intervention in Vanga to could be similar to the effectiveness of an intervention in Yasa Bonga, due to their geographical closeness and similarities. Meanwhile, the Kasaï Orientale coordination has been targeted for future VC interventions due to persistent gHAT case reporting, however, it is slightly more uncertain how successful deployment of Tiny Targets might be in this coordination to reduce tsetse density, due to not having past vector control interventions with a long history of tsetse monitoring nearby; this is approximately 600 km away from Yasa Bonga. As such, we have slightly less certainty on VC effectiveness for Mulumba.

### Fundamental Objectives of the Policy Maker

To determine what value can be gained from VC monitoring in our AAM framework, we must have a clearly defined fundamental objective. The fundamental objective determines our priority when comparing strategies and allows us to quantify their success.

An objective we could consider is reaching the target of elimination of transmission (EoT). When modelling transmission, one can compute when the last transmission event (LTE) occurs and use this as an indicator for EoT. However, the LTE is an unobservable event in real-world settings, meaning that policymakers are unlikely to know when it has occurred Further discussion on these metrics of EoT can be read in Sutherland et al [29]. However, there are issues with reaching EoT being the fundamental objective. In particular, reaching EoT at all costs is not a likely viable strategy, due to the economic limitations on interventions. For example, the best thing to do to eliminate transmission may be to attempt to kill every single tsetse to ensure no further infection spreads, but this is neither economically feasible in the DRC nor needed to reach elimination of the parasite.

As a result, it is important to consider strategies that minimise both the societal costs of the disease and the costs associated with interventions aimed at reducing its transmission. Therefore, our fundamental objective was to maximise the net monetary benefit (NMB), i.e. the difference in economic costs (including those from disease burden) from our proposed strategy to the baseline strategy.

Without treatment, or when treatment is delayed, patients with gHAT progress through two disease stages and most eventually die. The burden of disease for a given health zone under a specific strategy was estimated by weighting the expected person-time spent in each disease stage by the corresponding disability weights and adding the expected years of life lost due to disease-related mortality. This provided an estimate of disability-adjusted life years (DALYs). DALYs can be used to assess the burden the disease is expected to cause amongst the population. We prefer the use of DALYs here to other statistics, such as reported deaths and case numbers, for various reasons; in particular, estimated DALYs overcome the variable detection efforts for gHAT and deaths occurring without being reported. Furthermore, DALYs are useful to be able to compare with other causes of burden. Although it is a non-straightforward exercise, it is possible to assess what the willingness to pay (WTP) to avert DALYs has been historically [30]—the amount of money a government or donor is willing to pay to prevent the equivalent of one year of life lost—or to estimate what future funding would be prepared to pay. By multiplying DALYs averted by the WTP value, we can convert the health benefits of a strategy into monetary terms.

For the full calculation of the NMB including our unit costs, please see S1 Text.

### gHAT transmission model

To evaluate the impact of different strategies on the DALYs and costs for a health zone, we make use of a mathematical transmission model. We used the previously developed dynamic gHAT model, recently presented in Antillon et al, [27], which takes tsetse biology into account, and is fully described in S1 Text. The human transmission dynamics in the model consider low and high-risk humans separately, describing progression from susceptible to exposed to infected (stage 1) to infected (stage 2) to recovered or death. A compartmental diagram of the model (Model 7) for both the tsetse and human populations can be found included in the model description in S1 Text.

For the modelling of the human population, we employed a stochastic, tau-leaping method, which allows us to take into account the inherent randomness in infection dynamics and only outputs integer numbers of infected people. Specifically, we use a version of the tau-leaping method that employs a hash-based matching pseudo-random number generation technique. This version of the gHAT model is described in Sunnucks et al. [31], and ensures that compared realisations of the model have the same “environmental randomness” by matching the random seed for each event draw at every time step. This gives us the benefits of the stochastic modelling, which captures the random behaviour of infection dynamics—especially near elimination—but in a reproducible manner, without artificially creating additional between-strategy uncertainty. In this study, we made several pairwise comparisons; this hash-based matching method will ensure that any differences in outcomes between strategies are due to changes to the intervention parameters and not due to random number draws.

The tsetse component of the model is divided into several compartments to account for the different stages of the tsetse life cycle. The first of these represents the pupal stage (*P_V_* ), into which individuals enter following larval deposition at a rate dependent on the natural population carrying capacity. From there, the other compartments are the teneral (not yet blood fed) and susceptible class *S_V_* , the non-teneral (blood fed) and susceptible class *G_V_* , the exposed classes *E*_1*V*_ , *E*_2*V*_ and *E*_3*V*_ and then the infected class *I_V_* , in which tsetse can transmit trypanosomes to hosts. Tsetse are more susceptible to becoming infective whilst still in their teneral stage, i.e. before their first blood meal, in what is referred to as the teneral phenomenon [32]. The system of ordinary differential equations (ODEs) that describe the tsetse population in the gHAT model can be found in S1 Text. We do not use tau leaping for the tsetse population due to their much higher transition rates between compartments, which would require taking much smaller time steps to accurately simulate with tau leaping, and the unknown tsetse population size, which makes using integer population values infeasible.

VC in the model, originally developed and described in Rock et al. [33], is represented by an additional death rate to the tsetse population in all compartments other than the pupal stage, since pupae are not able to fly, meaning they cannot be killed by Tiny Targets. The rate of this additional death due to VC is affected by *f_T_* (*t*), the probability that a tsetse is killed by a Tiny Target when it is seeking a blood meal at time *t*. This depends on the effectiveness of the intervention and is initially equal to a parameter we call *p*_targetdie_ before decaying with time based on the logistic equation, resetting to *p*_targetdie_ every time VC is redeployed. The formulation for this term *f_T_* (*t*), with 6-monthly Tiny Target deployment, can be seen in S1 Text. The VC intervention that we are modelling is a large-scale intervention across a large area rather than a targeted intervention, and so we assume that the death rate of tsetse due to VC scales linearly with the tsetse population. An example of this VC effectiveness *f_T_* (*t*) and the effect of VC on the tsetse population in the model can be seen in Figure B of S1 Text.

In our simulation VC affects the entire health zone, rather than some proportion of it. To determine if this was a reasonable assumption to make, we performed an additional analysis to statistically assess how much of the health zone is affected by VC when it is solely deployed along large rivers. This analysis was conducted using data from the Yasa Bonga health zone [11]. We found that, whilst a 5km buffer around the large rivers has been previously given as the estimated area protected by the Tiny Targets, our statistical analysis suggests that the protected area could be much larger than 5km, as there is not sufficient evidence to suggest an unequal reduction in case reporting, after the introduction of vector control, in and outside of the protected area. Therefore, we assumed that VC covers the entire health zone. This full analysis can be found in S1 Text.

The transmission model parameters used for these simulations match the health-zone-specific posteriors previously estimated by fitting the gHAT model to human case data from 2000–2020 for the two health zones, and fixed, location-independent parameters [27] (see S1 Text for further details). The original case data was from the WHO’s HAT Atlas [34]. Modelling assumptions and their justifications can be found in the SI of Antillon et al [27].

### Cost model

Intervention costs against gHAT can be grouped into a few specific categories, the first of which is the cost of doing active screening for gHAT cases across the DRC, which will continue until EoT (3 years of no cases reported). Secondly, the cost of doing VC must be considered, taking into account the costs of each tiny target, the cost of deploying these traps, and the cost of travelling to the region. The VC strategy implemented will determine the level of investment required for each of these components. Finally, there is also the cost of monitoring, which, like the cost of VC, will depend on the monitoring strategy we use. We note that passive screening and treatment also occur on top of active screening, but these efforts continue regardless of the level of gHAT, and so are not a cost that can be reduced by changing strategy.

Costs are denominated in 2022 USD, and unit costs can be found in S1 Text. The willingness to pay value is the amount of money that a policy-maker is willing to spend to prevent the loss of a DALY, which measures the disease burden and years of life lost due to disease. In this study, we used a default WTP value of $153/DALY, which is the midpoint of the lower and higher historical WTP values for the DRC (adjusted for inflation) [30, 35]. Other modelling studies have used WTP values for the DRC ranging from $0 to $2000 per DALY [27], and old WHO guidelines recommended one or three times the GDP per capita of the country [36], equating to values of $653 and $1959 respectively [37]. Results for other willingness to pay values can be found in section 1.6 of S1 Text.

### Strategies for comparison

In 2000–2022, we simulate historical levels of active and passive screening and no VC. As the WHO case and screening data is not available after 2022, for 2023–2026 we assume that active screening continued at the mean of the previous 5 years (2018–2022) for this period. This allows us to obtain initial conditions for future projections from the start of 2027. The future strategies we consider all include the same diagnostic algorithm, treatment of cases and level of passive and active screening. As with the 2022–2026 period, for 2027 we use the mean level of active screening based on 2018–2022. We set passive detection rates to be equal to those estimated for the end of the fitted period (2020). Active screening cessation occurs after no cases are detected for three years, with an additional follow-up year of active screening occurring in year 5 [27].

Firstly, we considered fixed VC strategies. In the first strategy, VC begins in 2027 and continues until either three consecutive years without reported cases have been observed (a criterion similar to that used for AS cessation) or the end of the simulation period (2056) is reached, whichever occurs first; and the third has no VC. In each strategy where VC occurs, targets were deployed every 6 months. In addition to these pre-determined strategies, we add an AAM strategy. This involves undertaking VC during 2027 and 2028, and monitoring the tsetse population over this time period with catch data, before making a decision on whether to continue VC based on what we learn from this data. The four strategies we consider can be seen in Table 1.

**Table 1.** The different strategies we consider against gHAT in this analysis. We always consider screening to be taking place.

| Strategy | VC during first two years | VC after first two years | Medical screening |
| --- | --- | --- | --- |
| No VC | No | No | Yes |
| VC (Continuous) | Yes | Yes | Yes |
| VC (2 years only) | Yes | No | Yes |
| VC (AAM) | Yes | Depends on monitoring | Yes |

For each strategy, we can calculate the net monetary benefit (NMB), a measurement of the benefit of each strategy over a default strategy that takes into account the burden of the disease and the economic costs of an intervention. A 3% yearly discount is additionally applied to these values.

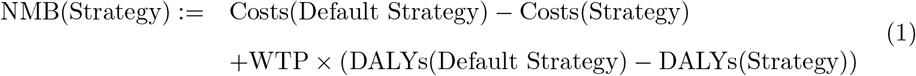

We considered the *No VC* strategy to be our default.

### Monitoring strategy

Pyramidal traps used for entomological monitoring [22] differ from Tiny Targets, which are used for VC. Pyramidal traps are designed to capture tsetse, whereas Tiny Targets are designed to kill tsetse, which often fly away from the Tiny Targets covered in insecticide. The locations of these traps are driven by factors specific to each field visit, with exploratory surveys to determine whether flies could be found in specific environments [11]. In our assessment of strategies, we assume that pyramidal trap monitoring is done before the initial VC intervention, which gives us an initial “T0” measurement to compare the later ones to. Once deployment of Tiny Targets has begun, we must consider when it is possible to do our monitoring. We considered monitoring which occurs immediately before any VC deployments. Given that the VC strategies considered involve biannual deployments, monitoring visits during the first two years were scheduled at 182, 365, and 547 days after the initial deployment.

In addition to deciding when to deploy our traps, we must also consider how many traps are deployed. The number of traps deployed has varied considerably in previous studies, ranging from 30 [16] to 1,229 [11], across areas of different sizes and at different times post-deployment. Increasing the number of traps is expected to reduce uncertainty in estimates of tsetse density, but this must be balanced against the associated increase in costs. This question of how many traps to deploy applies to both pre- and post-intervention measurements. The costs we used for undertaking monitoring can be found in S1 Text and comprise mainly of personnel time to conduct the monitoring and transport.

To appropriately interpret monitoring data, it is necessary to understand the distribution of trap catches — insect catch data are typically overdispersed, with many traps recording zero insects. [38]. Whilst there are a variety of other possible options, such as the zero-inflated Poisson, we assume a Poisson-gamma distribution, which is a Poisson-distributed random variable, with a gamma-distributed rate parameter. This can be re-parameterised into Poisson-gamma(*g, A*) where *g* is the mean and *A* is the overdispersion. This has expected value *g* and variance *g* + *Ag*^2^. It should be noted that when the overdispersion parameter becomes zero, the distribution is a regular Poisson, with rate parameter *g*. To estimate *A* without data from our exemplar health zones, we can use historical data from individual tsetse trap data from monitoring in Cote d’Ivoire [16] and the DRC [11]. Further information about how this was computed can be found in S1 Text.

We also need to know how to scale from the proportion of flies in our tsetse model to trap catch numbers. This scale parameter, *g*_0_ is the expected number of tsetse caught in a trap before the VC intervention.

### Active adaptive management

AAM is an approach designed to better support policy-making for a wide variety of applications, including combatting the spread of diseases. It involves learning about an action by doing it for an initial period and then using the information learned to refine our prior knowledge and better predict the best course of action after the initial period.

The problem is now established, with defined policy objectives, potential strategies, and an established monitoring plan. VC is then trialled for a short period, during which monitoring efforts take place. For our study of gHAT, this period will last two years, with the monitoring occurring as outlined in the previous section with pyramidal traps. At the end of this period, the data collected from monitoring efforts are then used to update prior beliefs on the effectiveness of the intervention. With these new updated beliefs, which should have less uncertainty than the prior beliefs, we can come back to our original problem, and use the objectives, possible strategies and modelling to ascertain what the next best course of action is. For our problem, this comes down to determining whether or not the expected NMB of continuing VC is higher than that of stopping VC.

The advantage of this over pre-defined/fixed strategies is that the final decision, after the initial intervention period, is better informed and more likely to be the optimal decision. An illustration of this can be seen in Figure 2. The trade-off, however, is that doing the intervention period itself could be sub-optimal, and there are additional costs associated with monitoring.

**Fig 2.**
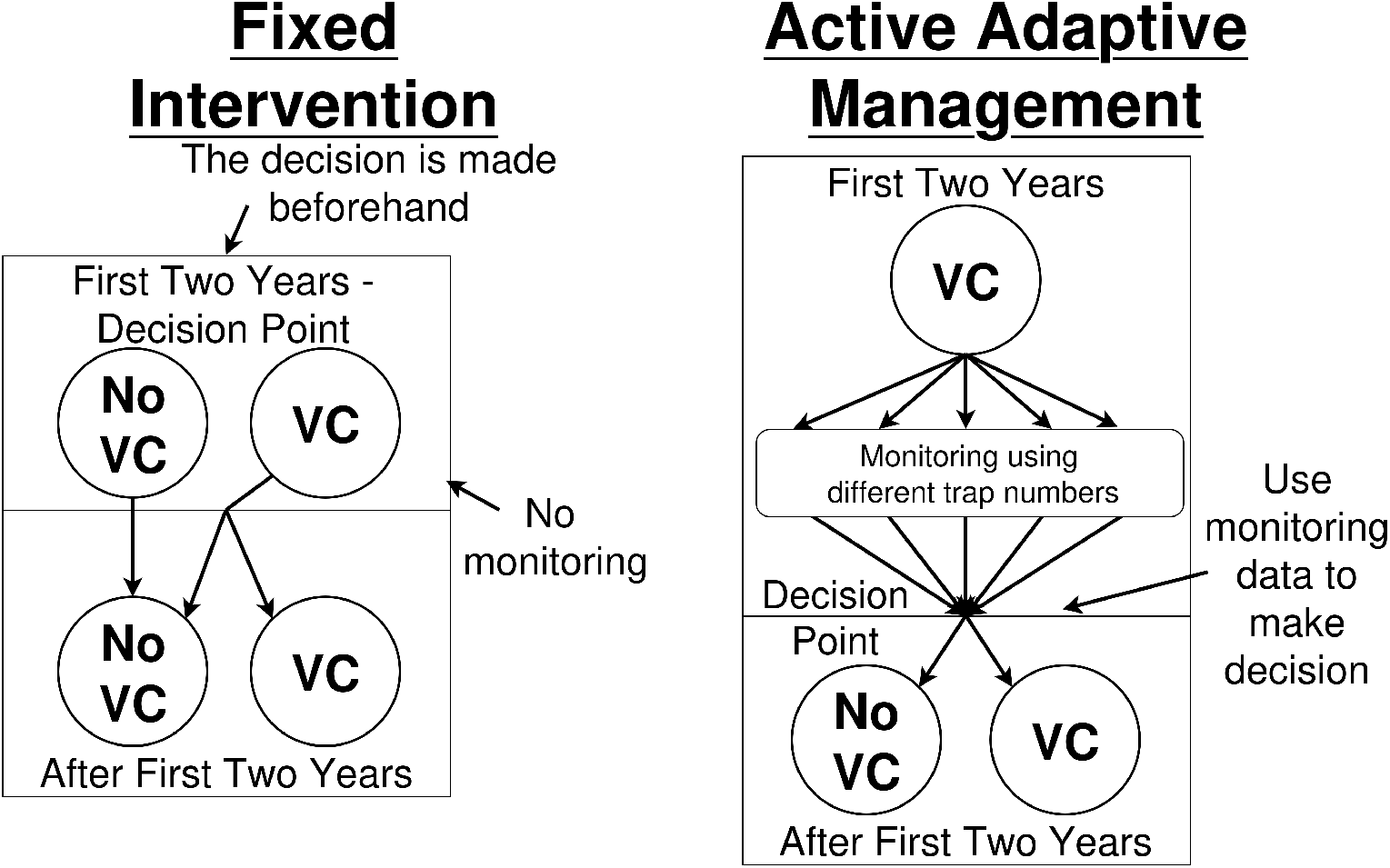
Illustration of the active adaptive management (AAM) framework for gHAT in comparison to fixed strategies and passive adaptive management. A fixed strategy is deciding on a specific intervention (i.e. no VC, VC for 2 years or continuous VC) and sticking with it for the full duration. For AAM, we undertake VC for the two-year period, and collect tsetse data in this time period to determine how effective VC has been, before then making a decision on whether to stop or continue VC.

### Our prior beliefs

We must set up prior beliefs on the parameters we wish to learn about through monitoring, using broad probability distributions to represent initial uncertainty. In this case, these parameters are those which relate specifically to VC and trapping.

The first of these is VC effectiveness, or the percentage reduction of the tsetse population after one year of VC. We represent this using a beta distribution, due to its property of being between 0 and 1, and the range of shapes the distribution can take. In the model implementation, this is converted to a probability of an individual tsetse hitting a Tiny Target and dying during its host-seeking phase of the feeding cycle, *p*_targetdie_. Our ODE model can be used to obtain the relation between percentage reduction and *p*_targetdie_, which can be seen in Figure C in S1 Text. It should be noted that we used two target deployments per year in our modelling, as is typically done in the DRC. The more frequently targets are deployed, the greater the reduction in tsetse abundance for a given value of *p*_targetdie_, because there is less time for target effectiveness to decline between deployments.

We consider two different prior beliefs on the effectiveness of VC, with the health zone of Mulumba in Kasäı Orientale having more uncertainty on the probable VC effectiveness than the health zone of Vanga in Bandundu Nord. This higher uncertainty in Kasäı Orientale is because the prior beliefs are based on previous VC interventions, which, in the DRC, have only occurred in the Bandundu Nord and Sud health zones. We called the more uncertain prior for Mulumba the “wide prior”, and the more certain prior for Vanga is called the “narrow prior”, reflecting the shape of their probability density functions.

Both of these priors have a modal VC effectiveness of 80% (i.e. tsetse populations are most likely to decline by 80% after one year of VC). This value is commonly assumed for VC effectiveness in the DRC [39] and has been observed in a range of field settings [11, 16, 17, 19]. The width of each prior distribution was informed by expert opinion. The prior distributions can be seen in Table 2.

**Table 2.** Table of the prior distributions for the parameters concerning tsetse monitoring and VC intervention data. VC effectiveness (%) is measured as the percentage reduction of tsetse after 1 year of Tiny Target deployments.

| Parameter | Prior Distribution | 95% CI | Mean |
| --- | --- | --- | --- |
| VC effectiveness (%) (narrow prior) | $\sim 100 \times \text{Beta}(4.5, 1.9)$ | [33.4, 95.9] | 70.6 |
| VC effectiveness (%) (wide prior) | $\sim 100 \times \text{Beta}(3.4, 1.8)$ | [25.0, 95.2] | 65.5 |
| Scale ($g_0$) | $\sim U(0, 10)$ | [0.25, 9.75] | 5 |
| Overdispersion ($A$) | $\sim \Gamma(124.7245, \frac{1}{0.046115})$ | [4.79, 6.80] | 5.75 |

For the overdispersion and scale parameters used to model the trapping process, we used uniform and gamma prior distributions, respectively (Table 2). The uniform prior for the scale parameter has an upper bound of 10 flies per trap before VC, a value that is substantially higher than those observed in previous monitoring studies in Ĉote d’Ivoire [16] and the DRC [11].” The overdispersion prior is based on this same previous monitoring data, and information on how it was obtained can be found in S1 Text.

### Using data and a likelihood function to update beliefs

At each monitoring time point, we expect to record the number of traps deployed and the tsetse catch for each trap. To update our uncertainty, we use the Poisson-gamma distribution of the data to then derive our likelihood function. This requires first defining the expected tsetse catch per trap at monitoring time *t_i_*, which is given by:

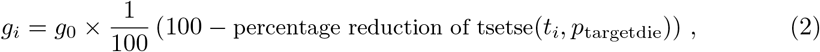

where the count from trap *j* at time *i* is

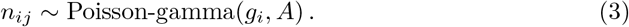

For any monitoring time, the percentage reduction in tsetse is determined by *p*_targetdie_. To avoid repeatedly solving the ODE system, we precalculated the relationship between *p*_targetdie_ and the corresponding percentage reduction at each of the four monitoring times.

From this, we can see the probability of observing *x* tsetse in a single trap (*j*) at time *i* is given by

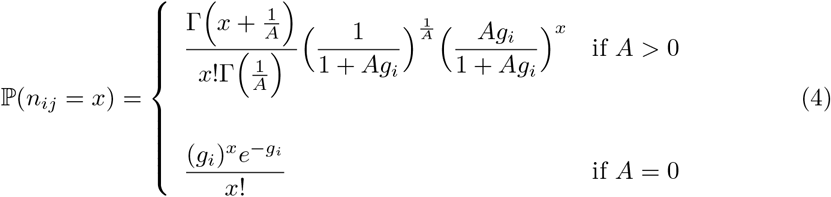

Noting that *x*! = Γ(*x* + 1), this gives us a log-likelihood of:

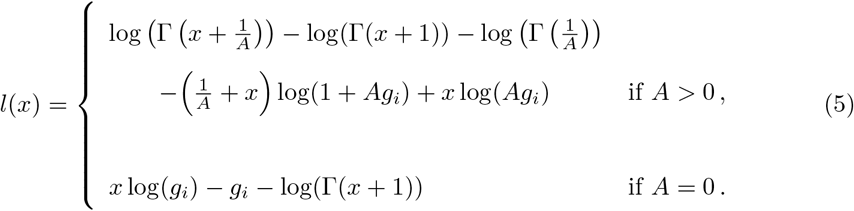

### Updating our beliefs

We have three different parameters per health zone for which we have priors—*A, g*_0_ and *p*_targetdie_. Because of the three-dimensional parameter space, it is not so simple to compute our posterior distribution for VC effectiveness. As such, we used an adaptive Metropolis-Hastings (AMH) method [40] which created a Markov chain that approaches the joint posterior distribution of the three parameters. The adaptive nature of this algorithm allows us to achieve a desirable level of mixing. The parameters *p*_targetdie_ and *g*_0_ are both explored in the log-space to ensure they stay positive and the acceptance rates are updated accordingly. The Matlab code for the AMH method can be found at osf.io/8q5ug.

By running this process, taking the chain and removing the initial burn in, we can obtain a strong approximation for the posterior distribution of *p*_targetdie_, from a set of tsetse catch data. Additionally, the longer we run the chain, the more accurate we become. These posterior distributions provide insight into the achieved tsetse reduction, and so, depending on how well we learn the reduction, provide insight into the effectiveness of monitoring strategies.

We can calculate the NMB for a range of *p*_targetdie_ values, and use a distribution (prior, *p*_0_(*p*_targetdie_) or posterior, *p*_1_(*p*_targetdie_)) to calculate an expected NMB of making a choice to continue or stop VC. Under the AAM strategy, a decision must be made after two years of monitoring. The decision rule is based on the posterior created from using the monitoring data:

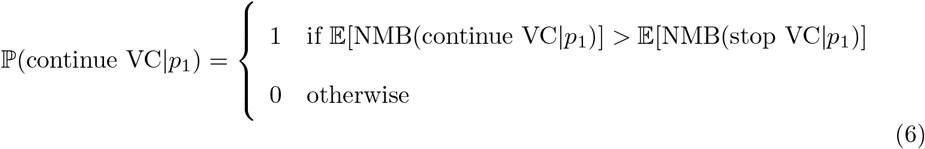

where

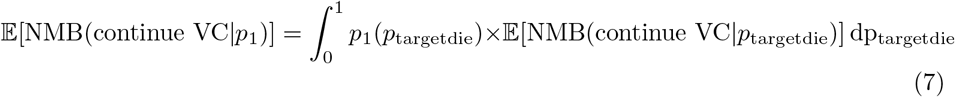

For a given true value of *p*_targetdie_, there is a large number of possible tsetse catch data sets. Let Ω be the set of all possible tsetse catch data, *p_A_* be our prior on overdispersion, *p_g_*_0_ by our prior on scale and L be our likelihood function stated above. Then, the probability of getting tsetse catch data *ω* is:

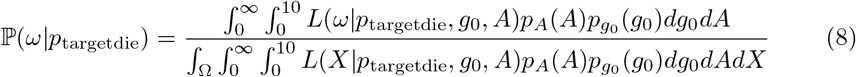

Then, defining *p*_1*,ω*_ as the posterior obtained from catch data *ω*, the probability that we would continue VC for a true underlying value of *p*_targetdie_ is:

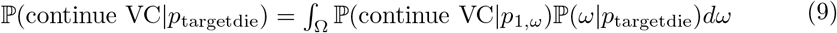

We note that, if the prior distribution is strong and poorly representative of the true value of *p*_targetdie_, and the number of traps per monitoring point is low, the probability of selecting a non-optimal decision increases, as the posterior distribution may not adequately reflect the true value. If the data is informative and the posterior reflects the true value well, the decision taken is more likely to be the optimal decision.

This decision rule allows us to compute the expected NMB for the AAM strategy for every value of *p*_targetdie_:

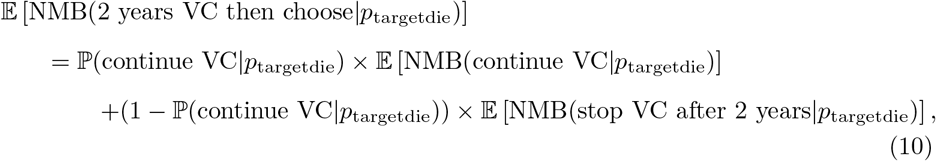

and, finally, we can determine the expected NMB of AAM over the original prior, given that we will learn the new information:

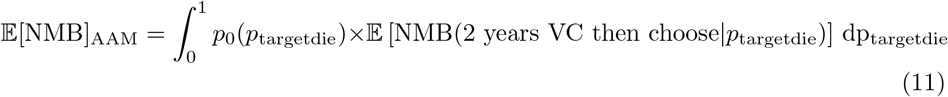

This then allows us to quantify the benefit of doing AAM, i.e. the difference in expected NMB between the best pre-defined strategy and doing AAM:

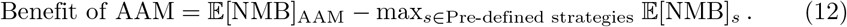

### Synthetic monitoring data

Updating the prior distributions requires the collection of monitoring data. As we are forecasting future VC and monitoring outcomes, we generated synthetic datasets representing possible future observations and use these to evaluate how decisions would be made under each scenario. To create these synthetic data, we use the Poisson-gamma probability distribution outlined in the calculation of our likelihood and selected parameter values for this distribution. For overdispersion and scale parameters, we drew these randomly from our priors, whilst many (135) values from across the entire range of possible *p*_targetdie_ values are considered.

We generated a large number (4050 — 30 per *p*_targetdie_ value) of possible data sets. For each dataset, we calculated the likelihood of observing these data for each value of *p*_targetdie_ and determined the resulting posterior distribution, which was then used to assess whether VC would be continued or stopped. This then allows us to estimate, for each possible value of *p*_targetdie_, the probability that the observed monitoring data would support continuation of VC

An example of this synthetic data can be seen in Figure 3, where the values caught in 100 traps were summed, using a *p*_targetdie_ value of 0.05 (equivalent to a 59.6% reduction in tsetse density after one year). Similar plots for *p*_targetdie_ of 0.02 (30% reduction) and 0.155 (95% reduction), as well as fewer traps, are shown in the S1 Text.

**Fig 3.**
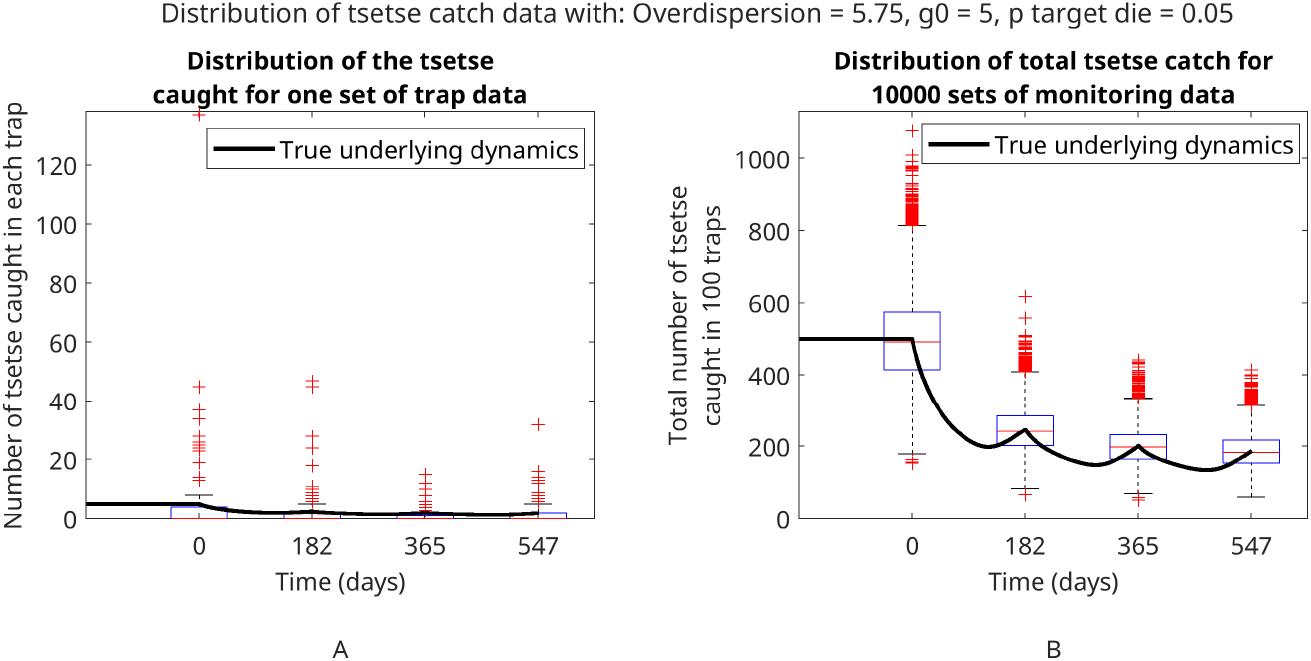
Example of possible monitoring outcomes given true underlying dynamics of the model. A shows the distribution of a single possible set of synthetic monitoring data and B shows the distribution of 10,000 sets of synthetic monitoring data, both with overdispersion *A* = 5.75, scale factor *g*_0_ = 5, and *p*_targetdie_ = 0.05 (a 59.6% reduction in tsetse after 1 year). In this example, 100 traps are used at each time point, with the individual trap counts displayed in A, and the sum over the individual traps displayed in B. The underlying model dynamics of the tsetse population over time can be seen as the solid line. The boxes represent the 25% and 75% quartiles of the data, the red line in the box represents the median of the data, and the whiskers represent the minimum and maximum of all non-outlier data. Red crosses represent outlier data, which is classified as being more than 1.5 times the interquartile range away from the box.

### Ethics statement

This study did not directly use any human case data and used previously published model parameterisation from another publicly available modelling article [27]. Ethics approval was granted by the University of Warwick Biomedical and Scientific Research Ethics Committee (application number BSREC 80/21-22) to use the previously collected DRC country HAT data, provided through the framework of the WHO HAT Atlas [34], for the secondary modelling analysis by Antillon et al. [27]. No new data collection took place within the scope of this modelling study.

### Data availability

Data cannot be shared publicly because they were aggregated from the World Health Organization’s HAT Atlas which is under the stewardship of the WHO. Data are available from the WHO (contact or visit https://www.who.int/trypanosomiasis_african/country/foci_AFRO/en/) for researchers who meet the criteria for access. Full model code is available at osf.io/8q5ug.

## Results

Figure 4 shows the NMB over a range of VC effectiveness values for our three passive adaptive management strategies and the AAM strategy using a WTP of $153/DALY. Continuous VC was sometimes the preferred (optimal?) strategy compared with the other two passive adaptive management strategies, with two possible exceptions: (1) when the VC is not very effective, it is often preferable either not to implement VC or to implement it only during the initial two-year period, rather than continue the intervention indefinitely; (2) when VC is highly effective, to the point where the probability of future gHAT transmission is greatly reduced after only two years of intervention, there is limited value in continuing to pay for VC after the initial two year period as a substantial impact has already occured. This secondary exception does, however, only occurs in health zones when the reduction in tsetse after one year is very close to 100%.

**Fig 4.**
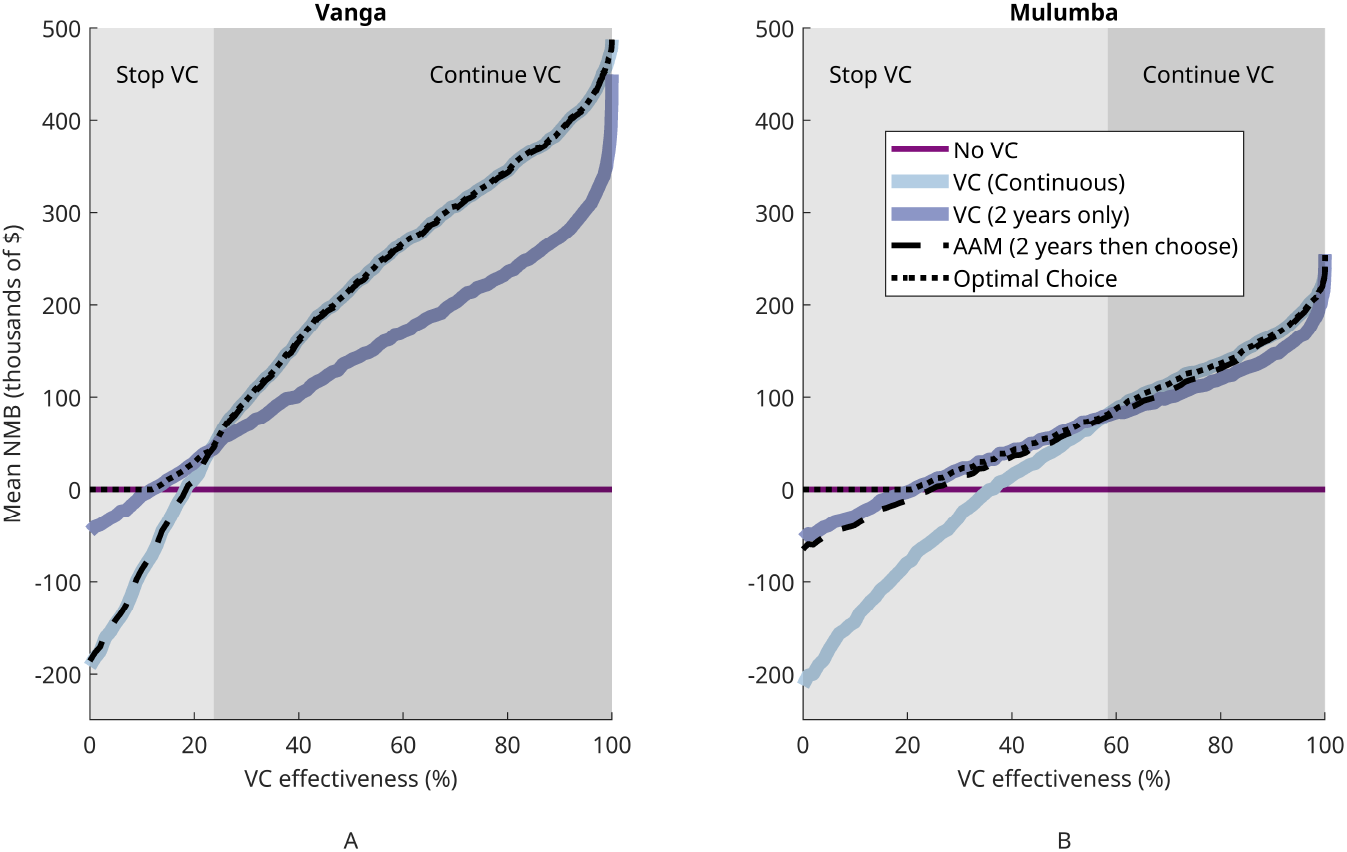
Net monetary benefit (NMB) for different vector control (VC) effectiveness values, with a willingness to pay (WTP) of $153/disability-adjusted life year (DALY), for two example health zones, Vanga (A) and Mulumba (B). Each of the coloured lines shows the NMB for a pre-defined strategy which does not use monitoring data to make a decision, whereas the black dashed line shows the NMB for the active adaptive management (AAM) strategy, where tsetse monitoring data using 96 traps is used to make a decision to continue or stop VC after two years. The regions where stopping and continuing VC are recommended have been shaded in light grey and grey, respectively.

Active adaptive management performed well in Mulumba, with outcomes close to the optimal strategy across all values of VC effectiveness. In Vanga, however, this was not the case: AAM resulted in outcomes very similar to those of the continuous VC strategy, with slightly higher NMB at low VC effectiveness and slightly lower NMB at high VC effectiveness

If the monitoring strategy provided perfect information (i.e. exact knowledge of *p*_targetdie_) after two years, AAM would recommend continuing VC when the continuous VC strategy has a higher NMB than the initial VC strategy for that value of *p*_targetdie_, and stopping VC otherwise. This represents the point where the light and dark blue lines cross in Figure 4. However, as the information gained is not perfect, we must determine which strategy has a higher expected value/NMB under the imperfect posterior. This may lead to decisions that are not optimal under the true value of VC effectiveness. We can then compare how different monitoring strategies affect how frequently the correct intervention decision is made. Figure 5 shows as we increase the number of traps used in our health zones, the closer our decision-making becomes to optimal.

**Fig 5.**
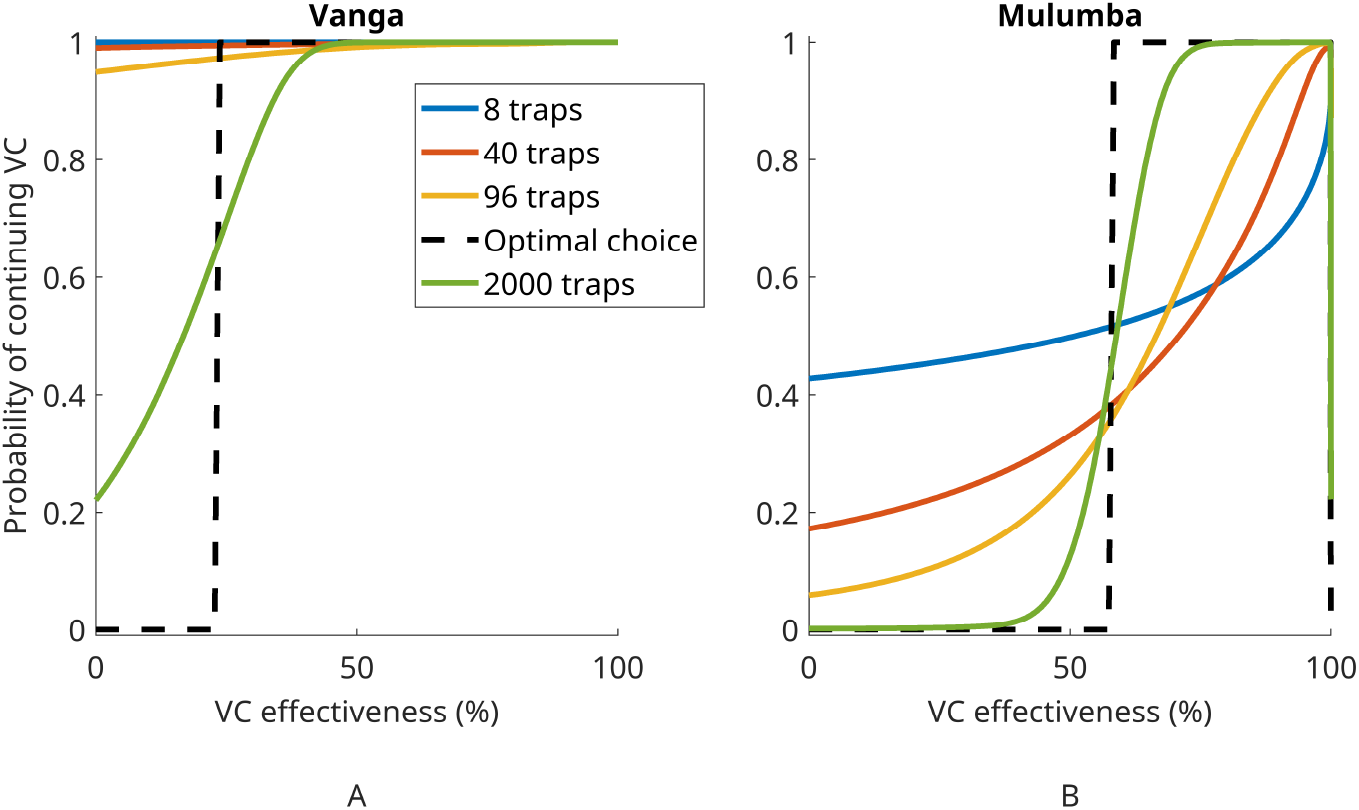
The probability of choosing to continue VC after 2 years based on monitoring data versus VC effectiveness (measured as the true reduction in tsetse population after 1 year). The optimal choice is shown as a dash black line. The coloured lines show the choice we would expect to make based on data from different numbers of traps to show the effect of increasing the monitoring efforts. These are produced using a willingness-to-pay (WTP) of $153 per disability-adjusted life year (DALY) averted. The unrealistic case of 2000 traps has been shown to demonstrate that as more data is collected, we get closer to more optimal decision making. The left-hand side (A) shows Vanga in Bandundu Nord and the right-hand side (B) shows Mulumba in Kasaï Oriental.

In Mulumba, we found that when using many traps, we usually stop VC for low effectiveness and continue VC for high effectiveness, as desired. For intermediate values of effectiveness, however, continuing and stopping VC are both likely outcomes. This is to be expected, as at this boundary point stochastic variation in the data makes it difficult to determine which side of the optimal decision boundary the system lies on If we had a lower value of overdispersion, or if we used even more traps, we would expect to lie closer to the optimal choice curve.

In Vanga, the results differed from those observed in Mulumba. Here, continuing VC is preferable to stopping VC for most VC effectiveness values, and stopping VC is only optimal when VC effectiveness is sufficiently low that the tsetse population is reduced by no more than 23% after one year of intervention Under our prior, we believe that is very unlikely, and so even with 96 traps, we are often not convinced that the VC effectiveness low enough to stop VC when it actually is. As a result, the AAM strategy leads to very little difference in decision-making, only occasionally swaying the decision-maker away from continuing VC in situations of low VC effectiveness.

The mean NMB of each strategy, and of doing AAM can then be calculated over our original prior. We can see in Table 3 the NMB of our various strategies. It is worth noting that all three VC strategies (the two fixed strategies and AAM) have positive expected NMB, indicating that they are preferable to no VC. This is not surprising as these health zones were unlikely to achieve elimination by 2030 without VC, and so have a moderate prevalence of gHAT. In particular, AAM had the highest mean NMB in Mulumba, where continuing or stopping VC resulted in similar outcomes, but not in Vanga, where continuing VC was considerably more beneficial than stopping. This is consistent with Figure 5, which showed that AAM had little impact on decision-making in Vanga but substantially improved decision-making in Mulumba.

**Table 3.** Table of mean net monetary benefits (NMBs) for various vector control (VC) strategies in each health zone, using a willingness-to-pay (WTP) of $153 per disability-adjusted life year (DALY) and 96 traps under active adaptive management (AAM).

|  | Vanga | Mulumba |
| --- | --- | --- |
| VC (continuous) | \$278,000 | \$93,300 |
| VC (Two years only) | \$187,000 | \$91,800 |
| AAM (2 years then choose) | \$276,000 | \$96,400 |

To specifically focus on the success of AAM, the metric ‘mean benefit of monitoring’ (see Equation 12) is useful and allows for the comparison of the AAM strategy with different numbers of monitoring traps. A non-negative mean benefit of monitoring implies that the cost of trapping is worth it, as the value gained in the resulting uncertainty reduction is more than the cost of undertaking the monitoring. In Figure 6, we can see the mean benefit of undertaking AAM as we vary the number of traps used.

**Fig 6.**
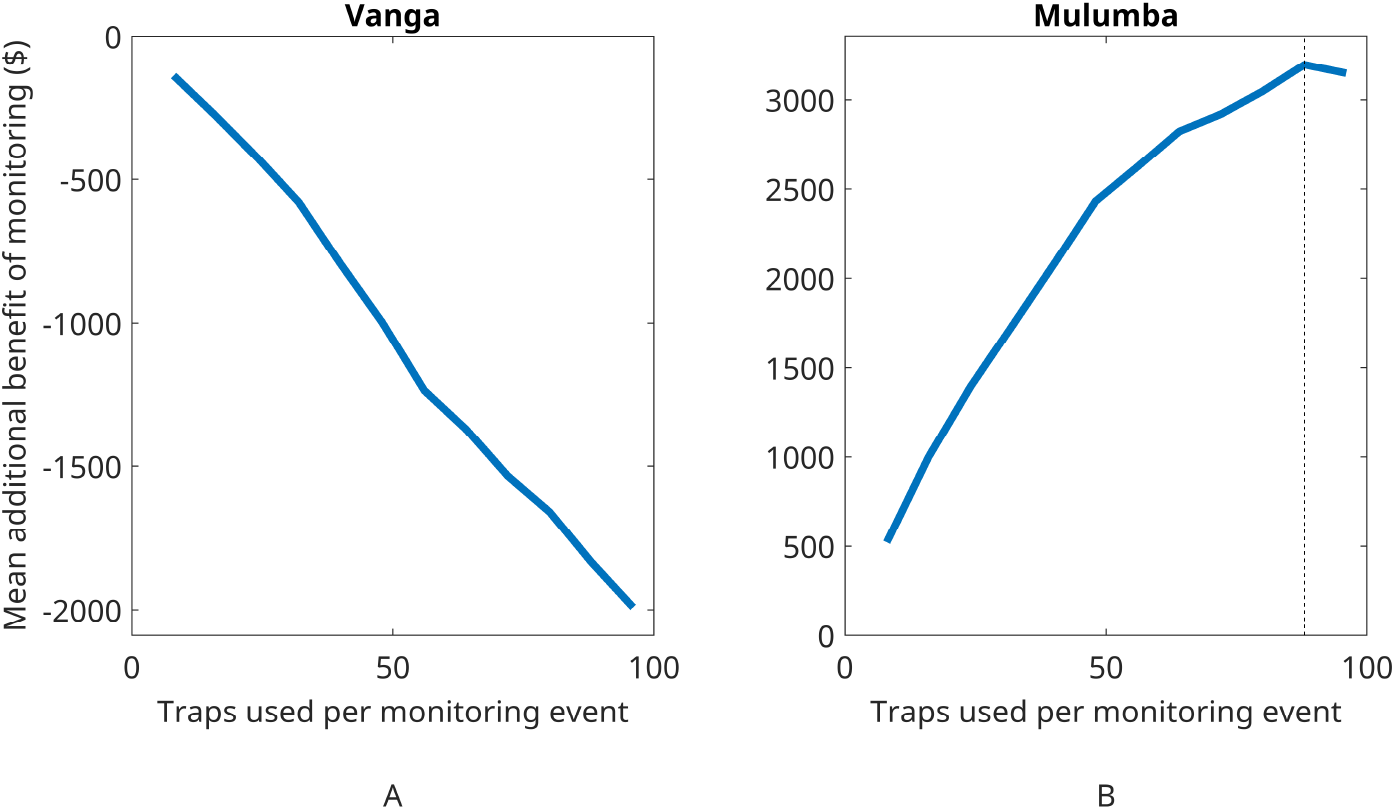
The mean additional benefit of undertaking the “do two years of vector control (VC) then choose” active adaptive management (AAM) strategy compared to conducting the best fixed strategy. The benefit is shown over a range of twice-per-year monitoring options with different numbers of traps to collect tsetse data. These are produced using a willingness-to-pay (WTP) of $ 153 per disability-adjusted life year (DALY) averted. The left-hand side (A) shows Vanga in Bandundu Nord, where benefit is negative and the right-hand side (B) shows Mulumba in Kasaï Oriental, where benefit is positive. N.B. The two y-axes have different scales.

For our intervention in Mulumba as seen in Figure 6, the slightly greater initial uncertainty and closer outcomes of stopping and continuing have resulted in AAM having a positive benefit, where the cost of monitoring is indeed worth it, and often is significantly exceeded by the value created by the monitoring process. In particular, monitoring with 88 traps maximised the value created. Increasing the number of traps beyond this point would eventually reduce the benefit, as monitoring costs increase while providing little additional information

Figure 6 illustrates a different scenario in Vanga. The combination of slightly higher initial certainty and the dominance of continuing VC over stopping VC has resulted in a negative benefit, which decreased further as the number of traps increased. Here, the information gained from data collection is unlikely to make a substantial impact on the decision-making and just increases the costs.

In summary, we extended the AAM framework to incorporate posterior updating for complex monitoring data, enabling assessment of the economic value generated by tsetse monitoring. By calculating expected NMB over the various prior distributions, we found that in both health zones, continuing VC was preferable over stopping VC after a two-year intervention when no information on the tsetse abundance was available. Then, under the AAM framework, we found that monitoring was cost-effective in the Mulumba health zone but not in the Vanga health zone. Finally, in Mulumba, we were able to find the monitoring strategies that resulted in the largest expected benefit of undertaking AAM by choosing to deploy 88 out of a possible 96 traps at each deployment time, although the full 96 traps would provide a very similar monetary benefit to only using 88.

## Discussion

In this study, we investigated how monitoring data can directly update prior beliefs on the effectiveness of VC. We considered how VC data can be used directly for decision making by laying out a framework under the active adaptive management methodology. This required the combination of Bayesian statistics and MCMC methods with epidemiological forecasting through the use of compartmental models.

By investigating the posterior distributions for a range of different monitoring strategies, we found that individual trap data does not provide noticeably more of an uncertainty reduction than aggregated data, due to the certainty we already have in our overdispersion prior (see S1 Text). Our overdispersion prior was obtained by extrapolating overdispersion from previous interventions in Côte d’Ivoire and the DRC. These interventions may not neccessarily be fully representative of future interventions in Vanga and Mulumba, and it may be reasonable to consider a wider range of overdispersion values. If this were the case and we had a more uncertain prior for overdispersion, individual trap catch data would improve model fitting compared with aggregate data, as they would contain more information about the level of overdispersion It is worth noting that individual trap-level data are generally more valuable than aggregate data, as the existing individual trap observations enabled us to define an informative prior for overdispersion. Therefore, we believe that having access to individual trap data is preferable and should generally be straightforward to provide as this is how the data is collected. However, the specific value of individual vs aggregated will depend on the extent to which information can be extrapolated between locations and on the specific modelling objective. Other modelling approaches may also benefit more from individual-level data than the approach presented here. We also, unsurprisingly, found that increasing the number of traps reduced uncertainty.

Taking this concept of posterior updating, we extended the AAM framework to deal with this more complex data, being able to assess the economic value gained from tsetse monitoring. Beyond the two health zones we considered, we can consider what types of location would find monitoring to be beneficial under this framework. Applying this analysis to other settings or even for different strategy combinations should be easy, as the underlying analysis would be very similar. Locations with low prevalence of gHAT are unlikely to benefit substantially from monitoring efforts, as the VC would likely interrupt any remaining transmission during the monitoring period. With moderate levels of prevalence, it then becomes possibly useful to help determine whether or not VC should continue. For locations with high prevalence, it would likely only be useful if we had a high level of uncertainty on VC effectiveness beforehand, as if we were already quite confident on VC effectiveness not being low, then we might expect to always want to continue VC.

This is the first time anyone has explored the utilisation of VC monitoring data for making strategy decisions. Other modelling for gHAT has used fitted epidemiological data coupled with model projections for deision making but has generally considered fixed strategies which do not factor in collection of new data to modify the strategy, except for stopping all vertical interventions once a pre-determined number of years of no cases has been met [41].

This is also novel for active adaptive management in epidemiology. This study considers the cost of data collection itself, and varying the amount of data collected. Rather, in previous AAM work, the question is often whether or not it is worth undergoing an intervention of unknown effectiveness for a brief period in order to learn the effectiveness better [24]. Additionally, this work presents a way to do AAM with data which does not fit into the system of neat updating method with conjugate priors [25] (e.g. a beta prior with binomial data), rather than more complex priors and data format. This work outlines methods to do the AAM approach with MCMC Bayesian updating, opening up the possibilities for using AAM for different systems where neat conjugate priors do not exist. This could be for learning about other vector impact of interventions or many other monitoring situations.

We are considering tsetse species (Palpalis group spp.) associated with riverine forest habitats. Unlike Bandundu, Kasai is a more typical savannah environment: it is flatter, has fewer small streams, scarcer vegetation, and probably lower wildlife availability (and therefore fewer potential hosts for tsetse). Overall, this likely represents a less suitable environment for tsetse persistence. However, there may still be pockets of more suitable habitat where tsetse populations can concentrate. These patches would be separated by less suitable areas, potentially limiting tsetse movement and increasing spatial heterogeneity. Such conditions could increase uncertainty and contribute to higher overdispersion when trap catches from multiple locations are combined. Another direction for future studies could be to further extend this framework for its application to gHAT in the DRC. Different regions could be assessed, with the possibility of considering how efforts in one region affect another in terms of cost reductions, how tsetse habitat might vary in different regions, and how data collected in one region may affect prior beliefs on VC effectiveness in other regions.

Beyond the gHAT in the DRC, the methods outlined in this study could be extended to further diseases. Other NTDs that have VC deployed to combat their spread could use a slightly adapted version of this framework to fit their modelling of the disease and VC. Additionally, the AAM procedure can be extended to further diseases with slow case reporting and other possible implementation indicators with complex data.

### Limitations

This study made various assumptions in its analysis. As mentioned previously, tsetse are *k* strategists, who have population sizes that tend towards carrying capacity. However, in reality, there are environmental fluctuations to this carrying capacity, predominantly the fluctuations between dry and wet seasons. This could affect the exhibited reductions of tsetse, and so future work could consider how these fluctuations may affect monitoring.

We also note that in our modelling we have assumed homogeneous mixing between flies and people across the health zone. In reality, the coverage of VC will differ spatially. Smaller geographical units of health areas could be used to capture these differences, although it is worth noting that models at HZ level have been shown to capture the trends in case reporting for the DRC well. Additionally, we analysed VC coverage in S1 Text for the Yasa Bonga HZ, and found it would not be unreasonable to assume the effects of VC can be applied across the whole HZ. However, in HZs with less extensive river coverage this may not be reasonable, and so different spatial scales could improve this modelling.

In this study, we assumed that VC would consist of two deployments per year. This frequency is typical of VC programmes in the DRC, where the lower observed effectiveness of Tiny Targets makes repeated deployments necessary to reduce the risk of tsetse population resurgence. This frequency is typical of VC programmes in the DRC, where high rates of tsetse reinvasion from neighbouring untreated areas make repeated Tiny Target deployments necessary to sustain the suppression of tsetse populations. This reinvasion of tsetse was not modelled in this work. Additionally, we assumed a 2-year initial intervention period and set monitoring time points to occur only before each VC deployment. These choices were made based on practical implementation considerations, informed by coauthors with on-the-ground experience. Future work could evaluate how changes in the number of VC deployments, the initial duration of VC, and the timing of monitoring affect the resulting strategies and AAM procedure.

Furthermore, future work could evaluate the effects of incorporating medical strategies, including improvements to passive screening, increased active screening, and the introduction of new drugs. Insight from experts on these medical strategies and how they could affect the overall intervention would aid future work.

The objective of elimination of transmission was not directly considered in this study, which instead focused on the more concrete objective of NMB. Expert opinion could allow for the consideration of both of these factors and how various outcomes compare to each other [42]. Additionally, a risk-neutral approach was taken to decision-making, using the expected NMBs. Future work could consider a more risk-averse decision-making process.

## Conclusion

In this study, we investigated how monitoring data on the tsetse fly population can be used to support decision-making for VC strategies to control gHAT. Sometimes, the additional data will not alter the choice indicated by epidemiological evidence or model predictions, whereas in other cases it will lead to a change in strategy. The AAM methodology estimates the frequency with which additional data is expected to alter decisions and quantifies its economic value.

This work has demonstrated that monitoring efforts can provide valuable information for a policy-maker, and that investment in such activities may be justified. More broadly, we have developed a framework that can be applied to a wide range of diseases and interventions involving complex data collection. This could help justify funding for future data collection when the resulting information has the potential to improve decision-making and support disease control efforts.

## Acknowledgments

Thanks to all of the HAT MEPP team and collaborators for the ongoing discussions, which inspired the writing of this paper and for the development of the code that was used to generate the results presented here. The authors would like to thank Dr Erick Mwamba Miaka and the WHO HAT Atlas team for access to historical data from the DRC which allowed for the original gHAT model parameterisation in Antillon et al [27]. The authors would like to thank Professor Simon Spencer for assisting with the MCMC algorithm. For the purpose of open access, the authors have applied a Creative Commons Attribution (CC-BY) licence to any Author Accepted Manuscript version arising from this submission.

## Author contributions

- Conceptualisation KR RS ED
- Methodology RS
- Software RS
- Formal Analysis RS
- Investigation RS
- Data Curation IT EM
- Writing - Original Draft RS
- Writing Review and editing All authors
- Visualisation RS
- Supervision KR ED IT
- Project administration KR ED
- Funding acquisition All authors

## Rights retention statement

For the purpose of open access, the author has applied a Creative Commons Attribution (CC-BY) licence to any Author Accepted Manuscript version arising from this submission.

## References

1. World Health Organization. Human African trypanosomiasis (sleeping sickness);. Available from: https://www.who.int/data/gho/data/themes/topics/human-african-trypanosomiasis.

2. World Health Organization. Control and surveillance of human African trypanosomiasis: WHO TRS N984;. Available from: https://www.who.int/publications/i/item/WHO-TRS-984.

3. World Health Organization. Human African trypanosomiasis (sleeping sickness);. Available from: https://www.who.int/news-room/fact-sheets/detail/trypanosomiasis-human-african-(sleeping-sickness).

4. Vreysen MJB, Seck MT, Sall B, Bouyer J. Tsetse flies: Their biology and control using area-wide integrated pest management approaches. Journal of Invertebrate Pathology. 2013;112:S15–S25. 10.1016/j.jip.2012.07.026.

5. Rafferty JP. K-selected species. Encyclopedia Britannica;. Available from: https://www.britannica.com/science/K-selected-species.

6. Steverding D. The history of African trypanosomiasis. Parasit Vectors. 2008;1:3. 10.1186/1756-3305-1-3.

7. Lindner AK, Lejon V, Chappuis F, Seixas J, Kazumba L, Barrett MP, et al. New WHO guidelines for treatment of gambiense human African trypanosomiasis including fexinidazole: substantial changes for clinical practice. The Lancet Infectious Diseases. 2020;20(2):e38–e46.

8. World Health Organization. Ending the neglect to attain the Sustainable Development Goals: A road map for neglected tropical diseases 2021–2030;. Available from: https://www.who.int/publications/i/item/9789240010352.

9. World Health Organization. Criteria and procedures for the verification of elimination of transmission of;.

10. Rock KS, Torr SJ, Lumbala C, Keeling MJ. Quantitative evaluation of the strategy to eliminate human African trypanosomiasis in the Democratic Republic of Congo. Parasites & vectors. 2015;8(1):532.

11. Tirados I, Hope A, Selby R, Mpembele F, Miaka EM, Boelaert M, et al. Impact of tiny targets on Glossina fuscipes quanzensis, the primary vector of human African trypanosomiasis in the Democratic Republic of the Congo. PLOS Neglected Tropical Diseases. 2020;14(10):1–20. 10.1371/journal.pntd.0008270.

12. Shaw APM, Torr SJ, Waiswa C, Cecchi G, Wint GRW, Mattioli RC, et al. Estimating the costs of tsetse control options: An example for Uganda. Preventive Veterinary Medicine. 2013;110(3):290–303. 10.1016/j.prevetmed.2012.12.014.

13. Food, of the United Nations AO. LIVESTOCK SYSTEMS IN SUB-SAHARAN AFRICA;. Available from: https://www.fao.org/4/j1255e/j1255e05.htm.

14. Shaw APM, Tirados I, Mangwiro CTN, Esterhuizen J, Lehane MJ, Torr SJ, et al. Costs Of Using “Tiny Targets” to Control Glossina fuscipes fuscipes, a Vector of Gambiense Sleeping Sickness in Arua District of Uganda. PLOS Neglected Tropical Diseases. 2015;9(3):1–19. doi:10.1371/journal.pntd.0003624.

15. Hope A, Mugenyi A, Esterhuizen J, Tirados I, Cunningham L, Garrod G, et al. Scaling up of tsetse control to eliminate Gambian sleeping sickness in northern Uganda. PLOS Neglected Tropical Diseases. 2022;16(6):1–23. doi:10.1371/journal.pntd.0010222.

16. Kaba D, Djohan V, Berte D, TA BTD, Selby R, Kouadio KADM, et al. Use of vector control to protect people from sleeping sickness in the focus of Bonon (Côte d’Ivoire). PLOS Neglected Tropical Diseases. 2021;15(6):1–18. doi:10.1371/journal.pntd.0009404.

17. Mahamat MH, Peka M, Rayaisse JB, Rock KS, Toko MA, Darnas J, et al. Adding tsetse control to medical activities contributes to decreasing transmission of sleeping sickness in the Mandoul focus (Chad). PLOS Neglected Tropical Diseases. 2017;11(7):1–16. doi:10.1371/journal.pntd.0005792.

18. Courtin F, Camara M, Rayaisse JB, Kagbadouno M, Dama E, Camara O, et al. Reducing human-tsetse contact significantly enhances the efficacy of sleeping sickness active screening campaigns: a promising result in the context of elimination. PLoS neglected tropical diseases. 2015;9(8):e0003727.

19. Hope A, Mugenyi A, Esterhuizen J, Tirados I, Cunningham L, Garrod G, et al. Scaling up of tsetse control to eliminate Gambian sleeping sickness in northern Uganda. PLoS Neglected Tropical Diseases. 2022;16(6):e0010222.

20. Checchi F, Filipe JA, Haydon DT, Chandramohan D, Chappuis F. Estimates of the duration of the early and late stage of gambiense sleeping sickness. BMC infectious diseases. 2008;8(1):16.

21. Checchi F, Funk S, Chandramohan D, Haydon DT, Chappuis F. Updated estimate of the duration of the meningo-encephalitic stage in gambiense human African trypanosomiasis. BMC research notes. 2015;8(1):292.

22. JP G, J L. Le piege pyramidal a tsetse (Diptera: Glossinidae) pour la capture et la lutte. Essais comparatifs et description de nouveaux systemes de capture [The pyramidal trap for collecting and controlling tsetse flies (Diptera: Glossinidae). Comparative trials and description of new collecting technics]. Trop Med Parasitol. 1986;37:61.

23. on Gambiense Human African Trypanosomiasis NMCDG. Insights from quantitative and mathematical modelling on the proposed 2030 goal for gambiense human African trypanosomiasis (gHAT). Gates Open Research. 2020;3:1553.

24. Shea K, Tildesley MJ, Runge MC, Fonnesbeck CJ, Ferrari MJ. Adaptive Management and the Value of Information: Learning Via Intervention in Epidemiology. PLOS Biology. 2014;12(10):1–11. doi:10.1371/journal.pbio.1001970.

25. Atkins BD, Jewell CP, Runge MC, Ferrari MJ, Shea K, Probert WJM, et al. Anticipating future learning affects current control decisions: A comparison between passive and active adaptive management in an epidemiological setting. Journal of Theoretical Biology. 2020;506:110380. 10.1016/j.jtbi.2020.110380.

26. Wasserberg G, Osnas EE, Rolley RE, Samuel MD. Host culling as an adaptive management tool for chronic wasting disease in white-tailed deer: a modelling study. Journal of Applied Ecology. 2009;46(2):457–466. 10.1111/j.1365-2664.2008.01576.x.

27. Antillon M, Huang CI, Sutherland SA, Crump RE, Brown PE, Bessell PR, et al. Cost-effectiveness of end-game strategies against sleeping sickness across the Democratic Republic of Congo. medRxiv. 2024;doi:10.1101/2024.03.29.24305066.

28. Snijders R, Shaw APM, Selby R, Tirados I, Bessell PR, Fukinsia A, et al. The cost of sleeping sickness vector control in the Democratic Republic of the Congo. medRxiv. 2024;doi:10.1101/2024.02.02.24302172.

29. Sutherland SA, Madan JJ, Rock KS. Measuring elimination of gambiense human African trypanosomiasis: A comparison of deceptively different metrics. medRxiv. 2025; p. 2025–08.

30. Woods B, Revill P, Sculpher M, Claxton K. Country-Level Cost-Effectiveness Thresholds: Initial Estimates and the Need for Further Research. Value in Health. 2016;19(8):929–935. 10.1016/j.jval.2016.02.017.

31. Sunnucks R, Davis EL, Rock KS. Methods for Reproducible Comparison of Strategies in Stochastic Modelling. medRxiv. 2025;doi:10.1101/2025.10.09.25337145.

32. Haines LR. Examining the tsetse teneral phenomenon and permissiveness to trypanosome infection. Frontiers in Cellular and Infection Microbiology. 2013;3. doi:10.3389/fcimb.2013.00084.

33. Rock KS, Torr SJ, Lumbala C, Keeling MJ. Predicting the impact of intervention strategies for sleeping sickness in two high-endemicity health zones of the Democratic Republic of Congo. PLoS neglected tropical diseases. 2017;11(1):e0005162.

34. Franco JR, Cecchi G, Paone M, Diarra A, Grout L, Kadima Ebeja A, et al. The elimination of human African trypanosomiasis: Achievements in relation to WHO road map targets for 2020. PLoS neglected tropical diseases. 2022;16(1):e0010047.

35. in2013dollars com. Inflation Calculator;. Available from: https://www.in2013dollars.com/us/inflation/2016?endYear=2022&amount=1.

36. World Health Organization. World health report : 2002;. Available from: https://www.who.int/publications/i/item/9241562072.

37. Bank W. Congo [DRC] Data Commons;. Available from: https://datacommons.org/place/country/COD?utm_medium=explore&mprop=amount&popt=EconomicActivity&cpv=activitySource,GrossDomesticProduction&hl=en#.

38. Mugenyi A, Muhanguzi D, Hendrickx G, Nicolas G, Waiswa C, Torr S, et al. Spatial analysis of Gf fuscipes abundance in Uganda using Poisson and Zero-Inflated Poisson regression models. PLoS Neglected Tropical Diseases. 2021;15(12):e0009820.

39. Huang CI, Crump RE, Brown PE, Spencer SEF, Miaka EM, Shampa C, et al. Identifying regions for enhanced control of gambiense sleeping sickness in the Democratic Republic of Congo. Nature Communications. 2022;13:1448. 10.1038/s41467-022-29192-w.

40. Haario H, Saksman E, Tamminen J. An adaptive Metropolis algorithm. 2001;.

41. Davis CN, Rock KS, Antillón M, Miaka EM, Keeling MJ. Cost-effectiveness modelling to optimise active screening strategy for gambiense human African trypanosomiasis in endemic areas of the Democratic Republic of Congo. BMC medicine. 2021;19(1):86.

42. Antillon M, Huang CI, Rock KS, Tediosi F. Economic evaluation of disease elimination: an extension to the net-benefit framework and application to human African trypanosomiasis. Proceedings of the National Academy of Sciences. 2021;118(50):e2026797118.

