## Supplementary Appendix 1 for "Can non-routine data collection help decision-making for *gambiense* sleeping sickness? Using active adaptive management to assess the potential of vector control"

#### S1.1 The gHAT model

The *gambiense* human African trypanosomiasis (gHAT) model presented here has been extensively used by the Warwick "HAT MEPP" team for modelling transmission of the parasite *Trypanosoma brucei gambiense* between humans and tsetse in different settings. There have been numerous improvements to the model since its original publication; the version used here mimics that used by Antillon et al. [1] for modelling of gHAT in the Democratic Republic of Congo. Although all information is provided in this supplementary information (SI) and the code to make this analysis reproducible, for justification of modelling and intervention assumptions, the reader should refer to the SI of Antillon et al. [1]. A diagram of this compartmental model can be seen in Figure A.

In Table A, we can see a list of all the compartments in the model. This table begins with tsetse compartments before listing the human components. We note that the model code contains options for skin-only parasite infection [4] and for high-risk people to participate in active screening (AS) (or for low-risk people not to participate in AS), but we do not use these options in the present study.

##### S1.1.1 Parameterisation

The fitted parameters [1] seen in Table B generally correspond to assumed biological values that are not expected to vary in different locations. The fitted parameters in Table C come from model fitting work [1] done with MCMC to find posteriors. These posteriors and other information for our simulations do use real data pertaining to the past or planned activities of the Democratic Republic of Congo (DRC) health zones,

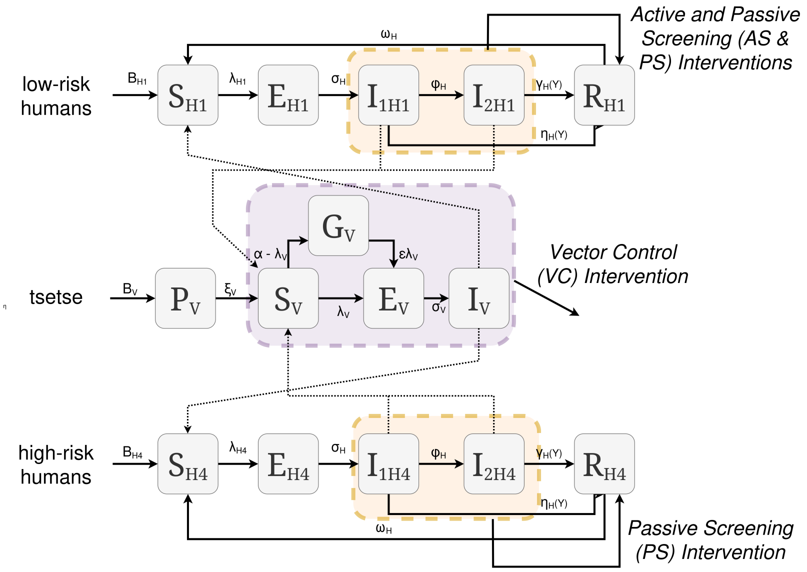


**Fig A.** Compartmental model of the tsetse and human populations modified from Sunnucks et al [2]. Also shown are the effects of passive screening, active screening and vector control interventions. We assume only low-risk people participate in active screening following Rock et al. [3].

| Compartment | Description |
|---|---|
| $P_V$ | The tsetse pupal class |
| $S_V$ | The tsetse susceptible, teneral class. |
| $E_{1V}$ | The tsetse exposed class (part 1) |
| $E_{2V}$ | The tsetse exposed class (part 2) |
| $E_{3V}$ | The tsetse exposed stage 3 class (part 3) |
| $I_V$ | The tsetse infectious class |
| $G_V$ | The tsetse susceptible, non-teneral class |
| $S_{Hi}$ | The human susceptible class |
| $E_{Hi}$ | The human exposed class |
| $I_{1Hi}$ | The human infectious stage 1 class |
| $I_{2Hi}$ | The human infectious stage 2 class |
| $R_{Hi}$ | The human recovered class |
| $S_A$ | The susceptible animal class |
| $E_A$ | The exposed animal class |
| $I_A$ | The infected animal class |

**Table A.** Compartments of the gHAT model. Tsetse are teneral before their first blood meal, when they have a higher susceptibility to becoming infected. We used the linear chain trick: having 3 exposed tsetse classes to create an Erlang-distributed extrinsic incubation period. Each human component has a low-risk ($i = 1$) and high-risk ($i = 4$) group.

although this data cannot be shared—instead a dummy data file will be provided with synthetic data. The tsetse-to-human relative density is calculated from $R_0$ [5]. The base probability of vector control (VC) killing a tsetse when it takes its bloodmeal, $p_{\text{targetdie}}$, is chosen to get the desired level of reduction in the tsetse population (which depends on the strategy).

| Notation | Description | Value |
|---|---|---|
| $N_H$ | Total human population size | 180,000 |
| $\mu_H$ | Natural human mortality rate | $5.4795 \times 10^{-5}$ days$^{-1}$ |
| $B_H$ | Total human birth rate | $= \mu_H N_H$ |
| $\sigma_H$ | Human incubation rate | 0.0833 days$^{-1}$ |
| $\varphi_H$ | Stage 1 to 2 progression rate | 0.0019 days$^{-1}$ |
| $\omega_H$ | Recovery rate | 0.006 days$^{-1}$ |
| $\xi_V$ | Pupal death rate | 0.037 days$^{-1}$ |
| $K$ | Pupal carrying capacity | $= 111.09 N_H$ |
| $\mathbf{P}$(pupating) | Probability of pupating | 0.75 |
| $\mu_V$ | Tsetse mortality rate | 0.03 days$^{-1}$ |
| $\sigma_V$ | Tsetse incubation rate | 0.034 days$^{-1}$ |
| $\alpha$ | Tsetse bite rate | 0.333 days$^{-1}$ |
| $B_V$ | Tsetse birth rate | 0.0744 days$^{-1}$ |
| $p_V$ | Probability tsetse infected per infectious bite | 0.065 |
| $\epsilon$ | Reduced non-teneral susceptibility factor | 0.05 |
| $f_H$ | Proportion of blood-meals on humans | 0.09 |
| Sens | Active Screening diagonostic sensitivity | 0.91 |
| $\mu_A$ | Natural animal mortality rate | 0.0014 |
| $\sigma_A$ | Animal incubation rate | 0.0833 |

**Table B.** Fixed parameters of the gHAT model. Adapted from Antillon et al [1]. We note that the birth rate parameter $B_V$ has been increased to lead to a higher possibility of the tsetse population heading to a non-zero equilibrium, as seen in health zones such as Yasa Bonga [6].

| Notation | Description | Percentiles of Prior Distribution [2.5, 50 & 97.5%] |
|---|---|---|
| $m_{\text{eff}}$ | Tsetse to human relative density | Calculated from $R_0$ |
| $R_0$ | Basic reproduction number (NGM approach) | [1.003, 1.069, 1.369] |
| $\eta_H^{\text{post}}$ | Stage 1 treatment rate | [4.59, 17.1, 42.9] $\times 10^{-5}$ |
| $\gamma_H^{\text{post}}$ | Stage 2 treatment rate | [2.33, 5.88, 12.0] $\times 10^{-3}$ |
| $\eta_{H_{\text{amp}}}$ | Relative improvement in passive screening stage 1 detection rate | [0.556, 2.893, 8.509] |
| $\gamma_{H_{\text{amp}}}$ | Relative improvement in passive screening stage 2 detection rate | [0.198, 1.137, 3.493] |
| $d_{\text{steep}}$ | Speed of improvement in passive screening detection rate | [0.761, 1.058, 1.424] |
| $d_{\text{change}}$ | Midpoint year for passive improvement | [2003.2, 2007.7, 2012.5] |
| $f_A$ | Proportion of blood meals on reservoir animals | [0.046, 0.5, 0.954] |
| $k_A$ | Relative size of animal reservoir population | [1.18, 18.3, 81.4] |

**Table C.** Fitted parameters of the gHAT model. Adapted from Antillon et al [1]

##### S1.1.2 Model equations

We implement a stochastic tau-leaping gHAT model for the human classes but a deterministic model for the tsetse population. In Table D we can see a list of each event that can occur within the stochastic tau-leaping gHAT model. The tau leap model takes discrete time steps of length 1 day, and draws Poisson random numbers to ascertain the number of each event that occurs. This version of the tau-leap model has had adjustments made to gain reproducible properties, where compared realisations share the same underlying randomness, only differing in their parameterisation [2]. For each of the events in our model, we will state the transition that occurs and the rate at which it occurs.

| Event | Transition | Rate |
|---|---|---|
| New Infection | $S_{Hi} \to E_{Hi}$ | $I_V \alpha m_{\text{eff}} f_i \frac{1}{N_{Hi}}$ |
| End Latency Period | $E_{Hi} \to I_{1Hi}$ | $\sigma_H p_{BS}$ |
| Die naturally in exposed state | $E_{Hi} \to S_{Hi}$ | $\mu_H$ |
| Progress to stage 2 of infection | $I_{1Hi} \to I_{2Hi}$ | $\varphi_H$ |
| Die naturally in stage 1 | $I_{1Hi} \to S_{Hi}$ | $\mu_H$ |
| Passive detection from stage 1 | $I_{1Hi} \to R_{Hi}$ | $\eta_H$ |
| Die naturally in stage 2 | $I_{2Hi} \to S_{Hi}$ | $\mu_H$ |
| Die naturally in recovered | $R_{Hi} \to S_{Hi}$ | $\mu_H$ |
| Lose immunity in recovered | $R_{Hi} \to S_H$ | $\omega_H$ |
| Passive detection or death from stage 2 | $I_{2Hi} \to R_{Hi}$ | $\gamma_H$ |
| New animal infection | $S_A \to E_A$ | $I_V \alpha m_{\text{eff}} f_A \frac{1}{N_A}$ |
| Animal becomes infectious | $E_A \to I_A$ | $\sigma_A$ |
| Exposed animal dies | $E_A \to S_A$ | $\mu_A$ |
| Infected animal dies | $I_A \to S_A$ | $\mu_A$ |

**Table D.** Events that can occur within the gHAT model, the transitions that occur, and the rate at which they occur per individual that can undergo the event.

**Passive detection:** The passive detection rates $\gamma_H$ and $\eta_H$ vary over time, and can be calculated as:

$$\gamma_H(Y) = \gamma_H^{\text{post}} \left[ 1 + \frac{\gamma_{H_{\text{amp}}}}{1 + \exp(-d_{\text{steep}}(Y - d_{\text{change}}))} \right]$$

$$\eta_H(Y) = \eta_H^{\text{post}} \left[ 1 + \frac{\gamma_{H_{\text{amp}}}}{1 + \exp(-d_{\text{steep}}(Y - d_{\text{change}}))} \right] \quad (1)$$

**Vector control:** The tsetse dynamics are simulated with an ODE model, as there are large numbers of events in a short time period. This is done with a Runge-Kutta approach to approximate the daily ODE tsetse dynamics [7]. The tsetse model is as follows, which can be seen in Equation system 2. The constants of 182.5 and 127.75 in the $f_T(t)$ term are chosen to obtain biannual behaviour that resets when targets are deployed, whereas the constant of 0.068 is chosen because it results in the believed decrease in effectiveness of Tiny Targets [8]. A plot of $f_T(t)$ and the effect of this on the tsetse population can be seen in Figure B. The relation of $p_{\text{targetdie}}$ vs percentage reduction after one year of VC can be seen in Figure C.

$$
\begin{aligned}
\frac{\mathrm{d}P_V}{\mathrm{d}t} &= B_V N_H - (\xi_V + \tfrac{P_V}{K})P_V \\
\frac{\mathrm{d}S_V}{\mathrm{d}t} &= \xi_V \mathbb{P}(\text{pupating})P_V - \alpha S_V - \mu_V S_V \\
\frac{\mathrm{d}E_{1V}}{\mathrm{d}t} &= \alpha\big(1 - f_T(t)\big)p_V\big(\textstyle\sum_i f_i \dfrac{(I_{1Hi} + I_{2Hi})}{N_{Hi}} + f_A \dfrac{I_A}{N_A}\big)(S_V + \varepsilon G_V) \\
&\qquad -(3\sigma_V + \mu_V + \alpha f_T(t))E_{1V} \\
\frac{\mathrm{d}E_{2V}}{\mathrm{d}t} &= 3\sigma_V E_{1V} - (3\sigma_V + \mu_V + \alpha f_T(t))E_{2V} \\
\frac{\mathrm{d}E_{3V}}{\mathrm{d}t} &= 3\sigma_V E_{2V} - (3\sigma_V + \mu_V + \alpha f_T(t))E_{3V} \\
\frac{\mathrm{d}I_V}{\mathrm{d}t} &= 3\sigma_V E_{3V} - (\mu_V + \alpha f_T(t))I_V \\
\frac{\mathrm{d}G_V}{\mathrm{d}t} &= \alpha(1 - f_T(t))\Big(1 - p_V\big(\textstyle\sum_i f_i \dfrac{(I_{1Hi} + I_{2Hi})}{N_{Hi}} + f_A \dfrac{I_A}{N_A}\big)\Big)S_V \\
&\qquad -\alpha\Big(f_T(t) + \big(1 - f_T(t)\big)p_V \varepsilon\big(\textstyle\sum_i f_i \dfrac{(I_{1Hi} + I_{2Hi})}{N_{Hi}} + f_A \dfrac{I_A}{N_A}\big)\Big)G_V \\
&\qquad -\mu_V G_V
\end{aligned}
$$

$$
f_T(t) = p_{\text{target die}}\left(1 - \frac{1}{1 + \exp(-0.068(\text{mod}(t, 182.5) - 127.75))}\right) \tag{2}
$$

**Active screening:** We model AS as occurring at the beginning of each year for the low-risk population. To do so, we utilise lists of the number of individuals that underwent AS in a given year. For future projections where these numbers are not known, we use the mean of the past 5 years to obtain a value. The number of infected individuals screened in stage 1 and stage 2, as well as the number of uninfected individuals screened, is then determined using a hypergeometric distribution. We then determine how many infectious screened individuals are detected, with a binomial distribution using the number of infectious individuals screened and the sensitivity of the test. Any positively tested infected individuals are then moved to the recovered class.

#### S1.2 Vector control coverage

VC is only conducted along the banks of major rivers in a given health zone. We wish to know whether or not this leads to an equal reduction in cases across the health zone, or if cases are not reduced as much far from the river. We will analyse the previous VC done in Yasa Bonga, where full VC was done along the region's major rivers from 2015 onwards. We have Yasa Bonga's gHAT screening data from 2006 to 2022, in addition to the knowledge of where screening was done. This screening data contains information on the location of each screening operation, the year in which the screening was conducted, and the number of cases detected. We can visualise this data by showing the locations where at least one case was detected, colouring them according to whether they were from before VC or after, as well as displaying a buffer around the rivers where VC took place. For this illustrative example, we will define before VC to be from 2006–2014, after VC to be from 2018–2022, and use a 10km wide buffer around the river. We exclude cases after 2014 and before 2018 due to the long incubation period of gHAT.

With this data, we can conduct some statistical tests on whether or not the VC has

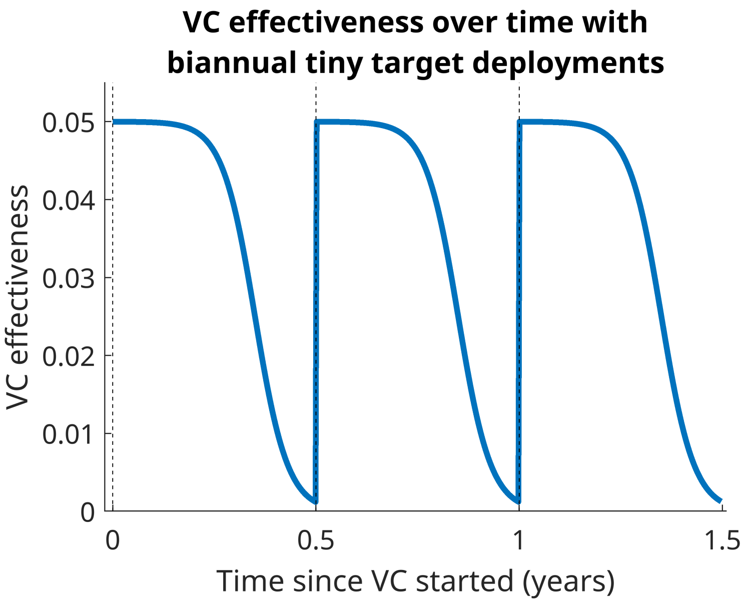


**(a)** Tiny Target effectiveness, which decays over time and is reset biannually.

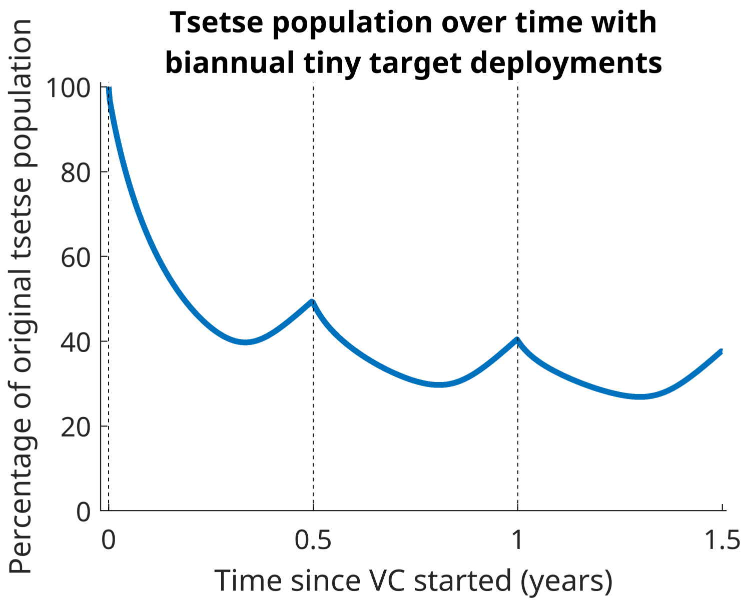


**(b)** Resulting reduction in tsetse population from the Tiny Targets

**Fig B.** Effect of Tiny Targets on the vector population, over a 1.5 year period. Biannual deployments of Tiny Targets are used (the times of which are shown with a dashed line), with a $p_{\text{target die}}$ value of 0.05.

affected the region around the river as much as the region outside the river. To do this, we must also ensure that the screening efforts are equal both before and after VC, which is something that cannot be guaranteed with the data set as it stands. To fix this, we can filter our data to only include points where AS was conducted both before and after VC. With this set up, we consider the following statistical test:

$$X_{\text{Before}} \sim \text{Binomial}(N_{\text{Before}}, p_{\text{Before}}) \qquad X_{\text{After}} \sim \text{Binomial}(N_{\text{After}}, p_{\text{After}})$$

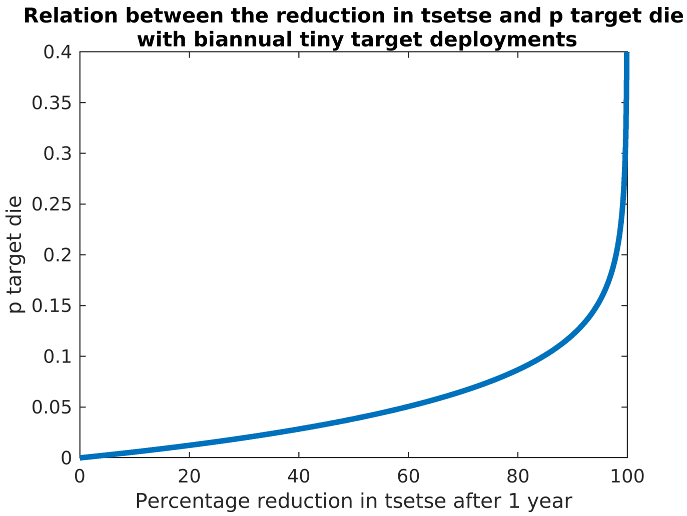


**Fig C.** The relation of $p_{\text{targetdie}}$ vs percentage reduction after 1 year of VC.

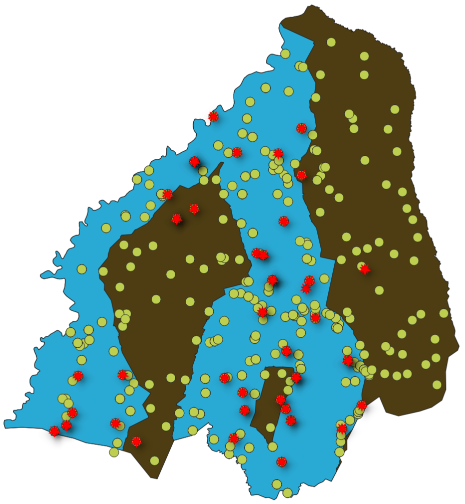

**Fig D.** Map of cases in Yasa Bonga, with cases pre-intervention marked as green circles, and cases post-intervention marked as red stars. A 10km buffer around the rivers where Tiny Targets were deployed can also be seen in blue.

Where, in time period $i \in \{\text{Before, After}\}$: 83

- $X_i$ is the total number of cases inside the river region, 84
- $N_i$ is the total number of cases across all of Yasa Bonga, 85
- $p_i$ is the probability a detected case is inside the river region. 86

Our null hypothesis is that the VC is equally effective inside the river region as it is 87 outside. This translates to the following hypothesis on the parameters $p_{\text{Before}}$ and $p_{\text{After}}$: 88

$$H_0 : p_{\text{Before}} = p_{\text{After}}$$

$$H_1 : p_{\text{Before}} > p_{\text{After}}$$

We can use a one-tailed proportion test to evaluate this, where our z-statistic is:

$$Z = \frac{\bar{p}_{\text{After}} - \bar{p}_{\text{Before}}}{\sqrt{\bar{p}(1-\bar{p})(\frac{1}{N_{\text{After}}} + \frac{1}{N_{\text{Before}}})}} \tag{3}$$

Where $\bar{p}_i$ is the observed proportion during time $i$, and $\bar{p}$ is the combined observed proportion. From this Z-value, we can obtain a p-value using the cumulative density of a normal distribution. A p-value of below 0.05 would indicate that their is sufficient evidence to reject the null hypothesis, whilst being above 0.05 would mean there is no evidence to reject our null hypothesis. Again, by defining before VC to be from 2006-2014, and after VC to be from 2018-2022, using a 10km buffer around the river, we obtain the data seen in Table E.

| | $X_i$ | $N_i$ | $\bar{p}_i$ |
|---|---|---|---|
| Before VC | 2252 | 2648 | 0.8505 |
| After VC | 35 | 41 | 0.8537 |

**Table E.** Observed cases inside the river region ($X_i$) and in the entire health zone ($N_i$), along with the observed proportion of the cases in the river region ($\bar{p}_i$), before and after vector control (VC).

This results in a Z-value of -0.0571 and a one-sided p-value of 0.52277, indicating that there is no evidence to reject the idea that VC is equally effective inside the river region as it is outside. We can conduct some sensitivity analysis however, varying what we consider to be pre and post-VC, as well as varying the size of the area around the river that we consider. The results of this can be seen in the tables below.

| | $X_i$ | $N_i$ | $\bar{p}_i$ | Z value | p value |
|---|---|---|---|---|---|
| Before (2006-2014) | 2252 | 2648 | 0.8505 | -0.0571 | 0.52277 |
| After (2018-2022) | 35 | 41 | 0.8537 | | |
| Before (2006-2014) | 2195 | 2583 | 0.8494 | 0.6201 | 0.26758 |
| After (2019-2022) | 16 | 20 | 0.8 | | |
| Before (2010-2014) | 460 | 523 | 0.8795 | 0.6129 | 0.26997 |
| After (2018-2022) | 33 | 39 | 0.8462 | | |
| Before (2010-2014) | 443 | 502 | 0.8825 | 1.1103 | 0.13344 |
| After (2019-2022) | 16 | 20 | 0.8 | | |

**Table F.** Results table for a 10km wide river region

| | $X_i$ | $N_i$ | $\bar{p}_i$ | Z value | p value |
|---|---|---|---|---|---|
| Before (2006-2014) | 782 | 2648 | 0.2953 | -0.3029 | 0.61903 |
| After (2018-2022) | 13 | 41 | 0.3171 | | |
| Before (2006-2014) | 764 | 2583 | 0.2958 | 0.4470 | 0.32743 |
| After (2019-2022) | 5 | 20 | 0.25 | | |
| Before (2010-2014) | 169 | 523 | 0.3231 | 0.5305 | 0.29789 |
| After (2018-2022) | 11 | 39 | 0.2821 | | |
| Before (2010-2014) | 164 | 502 | 0.3267 | 0.7188 | 0.23612 |
| After (2019-2022) | 5 | 20 | 0.25 | | |

**Table G.** Results table for a 5km wide river region

The p values found in the 5 and 10km tests are not sufficient to reject the claim that $p_{\text{Before}} = p_{\text{After}}$. It should also be noted, that the case where p-values were low were

| | $X_i$ | $N_i$ | $\bar{p_i}$ | Z value | p value |
|---|---|---|---|---|---|
| Before (2006-2014) | 2519 | 2648 | 0.9513 | 0.7192 | 0.23601 |
| After (2018-2022) | 38 | 41 | 0.9268 | | |
| Before (2006-2014) | 2454 | 2583 | 0.9501 | 2.0316 | 0.021096 |
| After (2019-2022) | 17 | 20 | 0.85 | | |
| Before (2010-2014) | 497 | 523 | 0.9503 | 0.7410 | 0.22935 |
| After (2018-2022) | 36 | 39 | 0.9231 | | |
| Before (2010-2014) | 476 | 502 | 0.9482 | 1.8803 | 0.030035 |
| After (2019-2022) | 17 | 20 | 0.85 | | |

**Table H.** Results table for a 15km wide river region

when the sample size was very low, with a large amount of successes, for which the normal approximation used in this test may not be appropriate.

We can analyse how these p values change with the width of the river region, to attempt to determine if we reach a point at which we can conclude that the infection dynamics are sufficiently different in and outside the region. However, as mentioned before, when the width of the region becomes too large or too small, the validity of the test can come into question due to having very few successes/failures. As such, we will exclude the distances for which the data is insufficient for this Z test. Specifically, we consider region widths that have at least 5 failures and successes in our binomial testing.

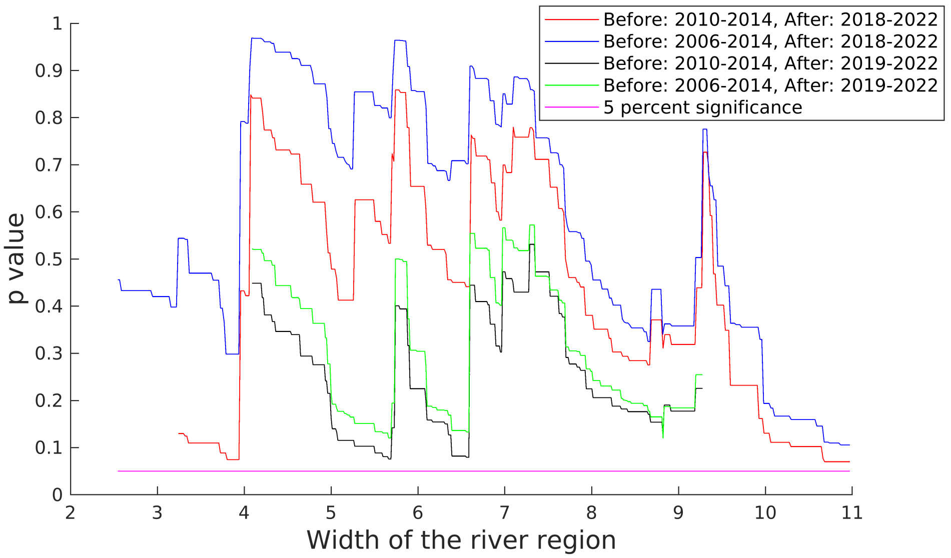


**Fig E.** p value of our hypothesis test for determining the effect of vector control away from the river, plotted against the width of the region around the river that is considered.

Now, only considering these regions where the test can be considered valid, we see that we do not reach the significance level of 0.05. Therefore, we lack evidence to reject the idea that the buffer around the river where VC is effective could be 10km wide (or even longer). Whilst we note that more data would help make a more informed decision, with these results, it is not unreasonable to assume that, for the sake of our modelling, in large-scale VC interventions, the VC fully covers the health zone when deployments cover sufficient amounts of the health zone.

#### S1.3 Cost function

We can calculate the disability-adjusted life years (DALYs) incurred from gHAT when we use a strategy. Using the model, the total person-years spent in each stage of infection can be calculated and the total number of deaths can be multiplied by the average years of life lost to gHAT to obtain the total person-years lost to death. With these and weights applied to the person-years spent infected, we can calculate the DALYs. The weights are 0.14 during the first stage of infection and 0.54 during the second stage [1]. The years lost to death is the life expectancy, 60.02, minus the average age at death of 26.63 [9], which, with the yearly discounting, becomes 21.54.

$$DALYS = 0.14 \times \text{PersonYears}_1 + 0.54 \times \text{PersonYears}_2 + 21.54 \times \text{Deaths}$$

With this, given a willingness to pay (WTP) for a DALY, we can calculate the 'cost' of the DALYs. We can consider several WTP values. Reasonable choices would be between 6.3 and 300, or possibly 653 and 1959 USD, corresponding to being between the lower and upper estimates of DRC WTP [10] converted from 2013 to 2022 USD [11], and then 1 and 3 times the DRC's gross domestic product (GDP) per capita [12], as recommended by the World Health Organization (WHO) [10].

Combining this with the costs of the intervention, we can calculate the net monetary benefit (NMB) of an intervention, using the no VC strategy as the comparative strategy. As such, to calculate the NMB in 2022 USD for a given year t, and a strategy S, we use the following comparative values:

- DALYs Averted$_{t,S}$ = DALYs in year t when doing the no VC strategy − DALYs in year t when doing strategy S
- Screening Difference$_{t,S}$ = $\mathbb{1}${Is doing AS in year t when doing strategy S} − $\mathbb{1}${Is doing AS in year t when doing the no VC strategy}
- Additional People Screeened$_{t,S}$ = People screened in year t when doing strategy S − People screened in year t when doing the no VC strategy
- Additional Infected Screeened$_{t,S}$ = Infectious people screened in year t when doing strategy S − Infectious people screened in year t when doing the no VC strategy
- VC$_{t,S}$ = $\mathbb{1}${Is doing VC in year t when doing strategy S}
- Monitoring$_{t,S}$ = The number of monitoring visits that do not coincide with VC deployments in year t when doing strategy S
- Traps$_{t,S}$ = Number of monitoring traps deployed during year t when doing strategy S

Noting that the fixed costs of passive screening cancel out, as these are unchanged between strategies, we get that for a year, $t$, the NMB for a strategy $S$ in 2022 USD is:

$$\begin{aligned}
NMB_t = {} & \text{WTP} * \text{DALYs Averted}_{t,S} \\
& - \text{Fixed AS Costs} * \text{Screening Difference}_{t,S} \\
& - \text{Testing Costs} * \text{Additional People Screened}_{t,S} \\
& - \text{Confirmation Cost} * \text{Additional Infected Screened}_{t,S} \\
& - (\text{Fixed VC Cost} + \text{VC cost per linear km} * \text{linear km of river}) * \text{VC}_{t,S} \\
& - \text{Fixed Monitoring Cost} * \text{Monitoring}_{t,S}
\end{aligned}$$

$$-\text{Cost per monitoring trap} * \text{Traps}_{t,S}$$

The linear km of rivers are specific to each health region and are determined from shape files for the DRC and its rivers. The number of monitoring traps deployed during the year t will vary for the AAM strategy and will be 0 otherwise. The rest of these are costs, each of which has a distribution taken from the literature. These are:

| Cost | Distribution | Reference |
|---|---|---|
| Fixed AS Costs | $\Gamma(56.18, 1412.88) + \Gamma(25.31, 747.18)$ | [1] |
| Testing Costs | $\Gamma(12.11, 0.1319)$ | [1] |
| Confirmation Cost | $\Gamma(8.47, 1.36)$ | [1] |
| Fixed VC Cost | $\Gamma(8.47, \frac{8.47}{9616.5*1.2194})$ | [13] |
| VC cost per linear km | $\Gamma(8.47, \frac{8.47*417}{43340.5*1.2194})$ | [13] |
| Fixed Monitoring Cost | $\Gamma(8.47, \frac{8.47}{5433*1.2194*0.5})$ | [13] |
| Cost per Monitoring Trap | $\Gamma(8.47, \frac{8.47*2472}{(4189+2610+1972)*1.2194})$ | [13] |

**Table I.** Cost Distributions for the calculation of net monetary benefit

Fixed AS Costs are the management costs of AS, plus the cost of an AS team, both taken from [1]. Testing costs are the cost of a CATT test, taken from [1] and confirmation cost is the cost of confirming a case of gHAT through microscopy, taken from [1]. We assume the fixed VC cost is the cost of travel, and we assume all other non-monitoring VC costs scale linearly with the extent (in linear km) of the VC. We assume the fixed monitoring cost is the cost of travel. We divide the cost listed in the article by two to get the cost per monitoring visit. These are only included during non-VC monitoring visits because otherwise, the team will already have to be out there to deploy VC during these monitoring visits. We then have all other monitoring costs scale linearly with the extent (in number of traps used) of the monitoring. All of the VC and monitoring costs are scaled by 1.2194 to account for inflation [11]. These values are taken from the Yasa Bonga VC costs [13] and scaled accordingly. As we only have one set of data to determine each of these costs, we take a gamma distributed cost, with the mean taken from these data and the 95% CI set to be 0.5 and 2 times the mean.

For this, we assumed that all other monitoring costs scale linearly with the extent (in number of traps used) of the monitoring. We also scale by 1.2194 to account for inflation. These values are taken from the Yasa Bonga VC costs [13] and scaled accordingly. As we only have 1 set of data to determine these costs (from Yasa Bonga), we take a gamma distributed cost, with the mean taken from these data and the 95% CI set to be 0.5 and 2 times the mean. Then the total NMB is the sum of the NMB of each year, with a 3 percent yearly discount applied.

#### S1.4 Overdispersion prior

We wish to have fairly well informed priors for our overdispersion and scale parameters. Unfortunately for our scale parameter it will be very difficult to gage because not only is there not much data one could use, but also one would expect it to be quite different for each location. As such, we use a uniform distribution for this that covers all possible values.

For the overdispersion parameter, we can attempt to fit this to data, as this is less likely to considerably vary from location to location. Additionally, we can use trapping data from each trapping time, rather than just the times before VC was deployed. To do this, we look at two sets of trapping data — one in Yasa Bonga in the DRC [6], and one in Cote d'Ivoire [14], and consider the scale and overdispersion at each trapping time. Firstly, we shall attempt to see if there is a clear relation between overdispersion and the scale parameter. To demonstrate this, we consider the relation between maximum likelihood estimates (MLEs) on scale and overdispersion from each trapping time. We plot them with a line of best fit in Figure F, and can also obtain the Pearson correlation coefficients. Unfortunately, a statistical test for independence cannot be carried out as we do not have a sufficient number of data points (only 20). We found that the correlation coefficient between all MLEs was 0.047857, between DRC MLEs was -0.14391 and between Cote d'Ivoire MLEs was 0.60998. Whilst the positive correlation does seem to imply that there could be a relation between overdispersion and scale in Cote d'Ivoire, when looking at the DRC there is a weak negative correlation, and when looking overall there is very little correlation. As such, we will assume there is no relation between scale and overdispersion for our tsetse data.

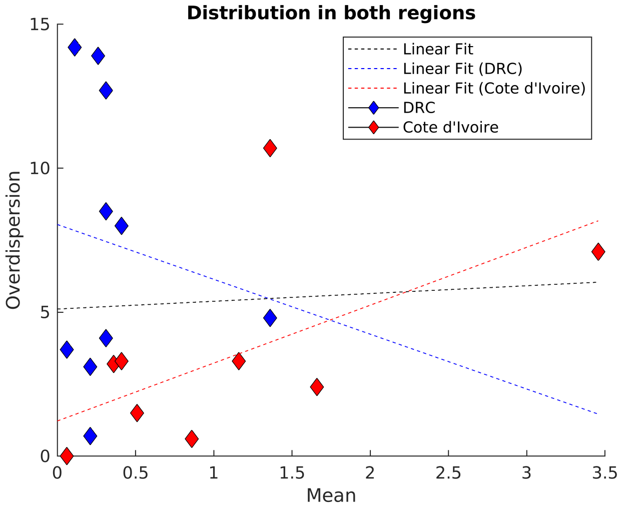


**Fig F.** Overdispersion and scale (mean individual tsetse catch) MLEs in historical trapping data in DRC and Cote d'Ivoire, with lines of best fit. Each point is a different monitoring time.

Now that we can be convinced that there is no (at least strong) relation between overdispersion and the scale parameter, we can go ahead to put a prior on overdispersion. We use the values of the likelihood function at each location to obtain a log-likelihood for a range of different overdispersion values from 0 to 20. These are then summed across the different trapping times, and locations, then converted into a likelihood function before being normalised to obtain a probability density function. In order to fit this prior, we then use the MATLAB function fitdist to fit this result to the gamma and lognormal distributions. The results of this can be seen in Figure G, and we found that the best fit was $\Gamma(124.7245, 0.046115)$, which we shall take to be our prior.

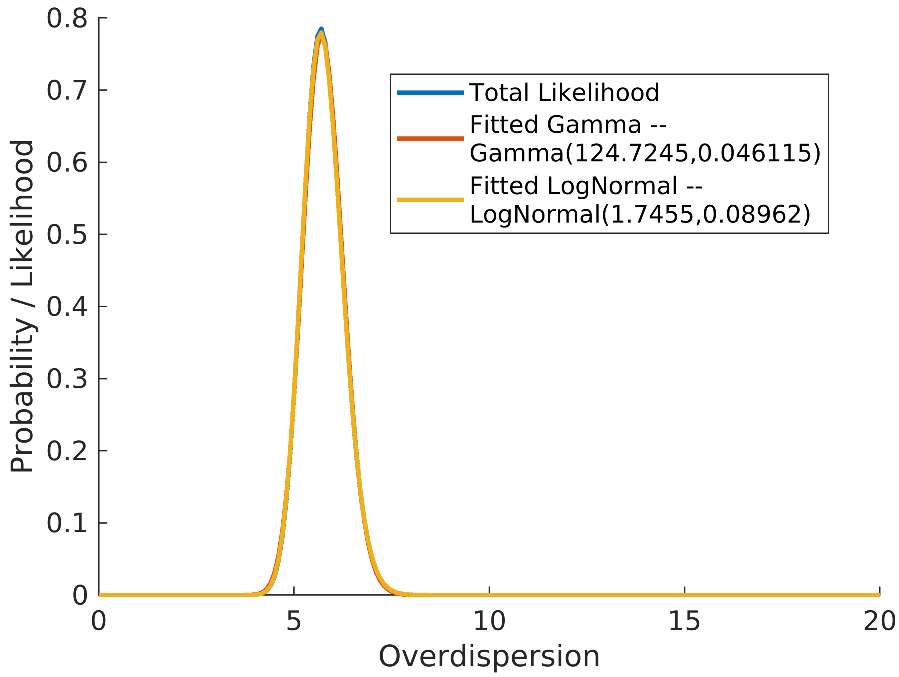


**Fig G.** Total likelihood function from all DRC and Cote d'Ivoire data, along with fitted priors (Gamma and LogNormal) on the overdispersion parameter.

#### S1.5 Posterior results

Before we do active adaptive management, we can seek to understand how the uncertainty changes for differing levels of monitoring. To do this, we will show how the posterior distribution on the effectiveness of VC converges to its true value for differing monitoring strategies. As an example, we can see what a typical posterior convergence looks like for one set of synthetic data in Figure H. Note how the posterior does not perfectly obtain the true value of effectiveness, due to remaining uncertainty and the randomly generated synthetic data possibly indicating a slightly different reduction in the tsetse caught.

We desire to learn specifically what type of data gives us the least error in our posterior convergence. We would expect that as we get more data (i.e. deploy more traps), the error of the posterior distribution will decrease. But there are other factors that could change this error. We can also ascertain how much of a benefit there is to having individual trap data, rather than just the summed values. i.e. finding out that beforehand we caught $0, 0, 0, 1, 1, ..., 2, 0, 3, 11, 0$ as opposed to learning we caught 250 tsetse with 150 traps. There should be some benefit, as individual data allows us to learn about the level of overdispersion present in the tsetse data that could cause additional unexpected variance. The benefit of individual data may also change as we vary the number of traps used.

To answer these questions about the effect on posteriors, we must quantify an error metric. We shall use a least-squares error metric on the posterior of the VC effectiveness, using the percentage reduction in tsetse after one year as a way to quantify effectiveness. We draw a 'true' value of percentage reduction $E^*$ from our prior as well as values of $g_0$ and $\alpha$ from their prior distributions. From these, we generate a set of data and then use

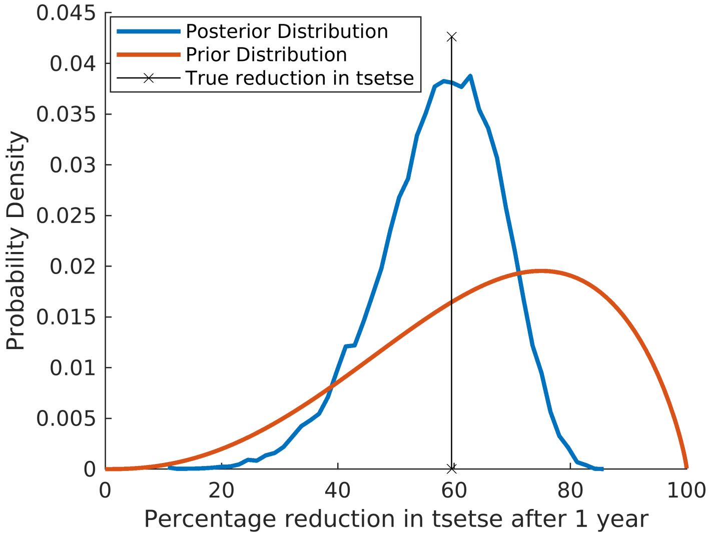


**Fig H.** Posterior convergence using only one set of synthetic data, of 100 traps deployed before as well as 182, 365 and 547 days in, with overdispersion $\alpha = 6$, scale factor $g_0 = 4$, and $p_{\text{target die}} = 0.05$.

this data to obtain a posterior distribution with probability density $f(x)$. Then,

$$Error = \int_0^{100} f(x)(x - E^*)^2 \mathrm{d}x.$$

We can then repeat this process multiple times, each time using new parameter draws and therefore new generated data, to obtain an average error value.

We shall compare the error as we vary the number of traps used, and whether or not we get individual or trapped data. We consider using between 10 and 25 traps at each time point, and will have monitoring done before the intervention as well as 182, 365 and 547 days into the intervention. We shall also demonstrate what would occur if we had a larger prior uncertainty on overdispersion, by considering a secondary overdispersion prior of:

$$\text{Overdispersion} \sim \Gamma(12, 0.4)$$

We can see that using individual trapping data results in a reduction in the error of posterior convergence if we have more initial uncertainty on overdispersion. This is likely because, under the current prior distribution, we are already quite certain as to the levels of overdispersion we expect, reducing the value of individual data. However, with more uncertainty on overdispersion, it becomes valuable to have individual trapping data to help us better learn the overdispersion. We also can see that, as expected, increasing the number of traps reduces our error in posterior convergence. Despite the fact that in our specific case, there is little benefit of individual trap data, we have used individual trap data for our fitting, in order to better demonstrate the method for a wider variety of use cases. We note that this does come at the expense of requiring more computation.

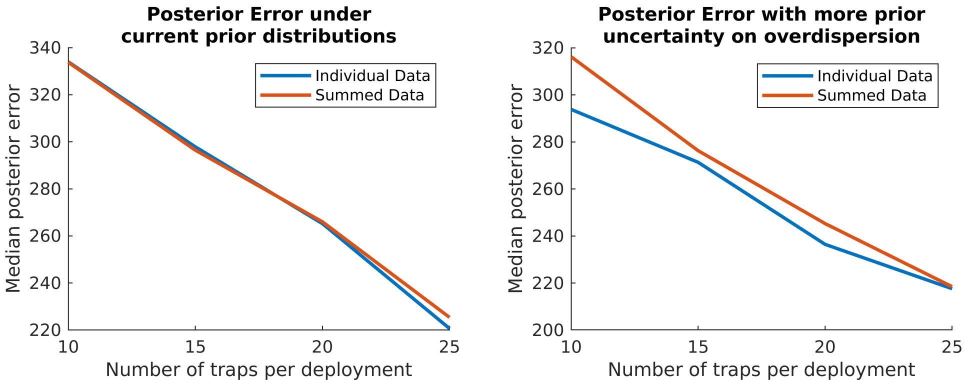


**(a)** Posterior error under the current overdispersion prior

**(b)** Posterior error a more uncertain overdispersion prior

**Fig I.** The median error in posterior convergence as we vary the number of traps and whether or not data is summed or individual. Median error taken over 100 potential data sets and corresponding posteriors.

### S1.6 Additional plots

#### S1.6.1 Additional synthetic data plots

In Figure J we can see the distribution of our synthetic data for a wider range of p target die values. We show values of 0.02, 0.05 and 0.155, which correspond to a 20%, 60% and 95% reduction in tsetse after 1 year, respectively.

#### S1.6.2 Additional willingness to pay (WTP) values

In Figures K and L we can see how the NMB and the mean additional benefit of monitoring change for Vanga and Mulumba as we alter the willingness to pay value from 153 to 653 and 1959, respectively. Here we see that in both cases, increasing the WTP makes VC continuation more worthwhile for low effectiveness values. As such, for both health zones, the benefit of doing monitoring ends up as negative and decreasing, as the additional information learned by undertaking monitoring is unlikely to change the decision away from continuing VC.

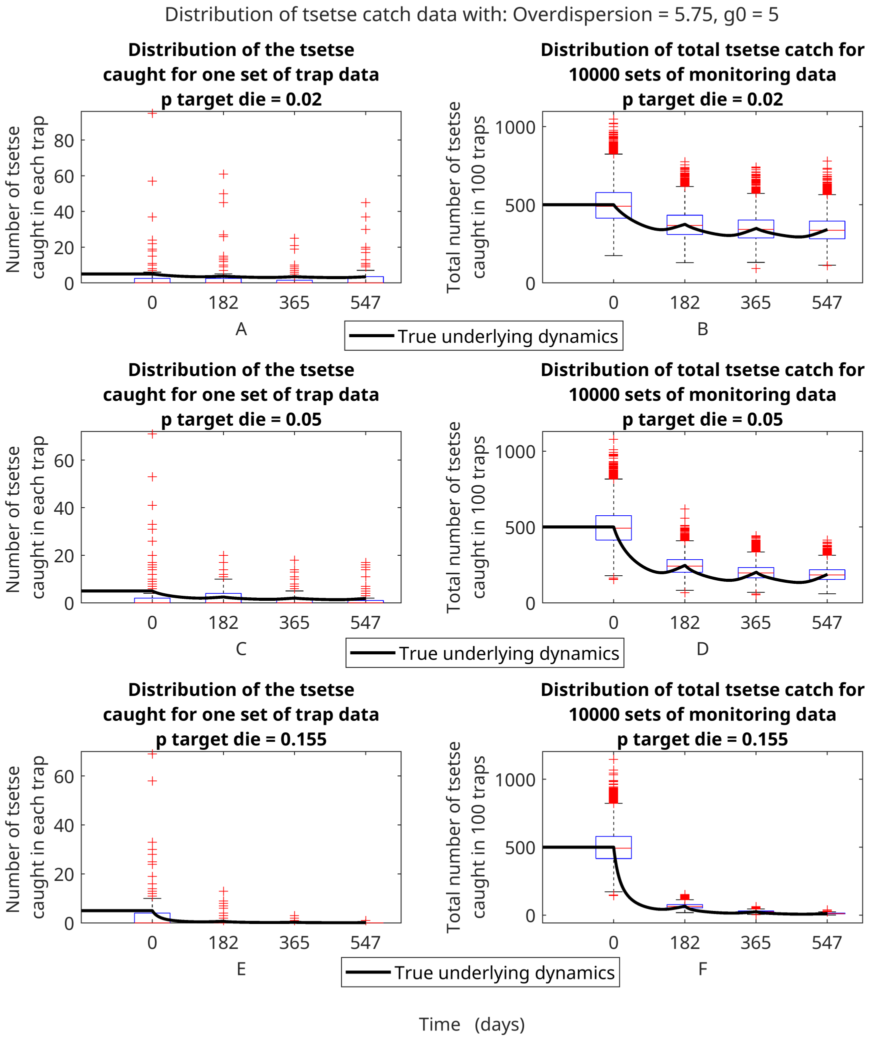


**Fig J.** Example of possible monitoring outcomes given true underlying dynamics of the model. A, C, and E (labels shown below each plot) show the distribution of a single possible set of synthetic monitoring data, and B, D, and F show the distribution of 10,000 sets of synthetic monitoring data, all with overdispersion $A = 5.75$ and scale factor $g_0 = 5$. For A and B, $p_{\text{targetdie}} = 0.02$ (a 30.2% reduction in tsetse after 1 year). For C and D, $p_{\text{targetdie}} = 0.05$ (a 59.6% reduction in tsetse after 1 year). For E and F, $p_{\text{targetdie}} = 0.155$ (a 95.0% reduction in tsetse after 1 year). In this example, 100 traps are used at each time point, with the individual trap counts displayed in A, and the sum over the individual traps displayed in B. The underlying model dynamics of the tsetse population over time can be seen as the solid line. The boxes represent the 25% and 75% percentiles of the data, the red line in the box represents the median of the data, and the whiskers represent the minimum and maximum of all non-outlier data. Red crosses represent outlier data, which is classified as being more than 1.5 times the interquartile range away from the box.

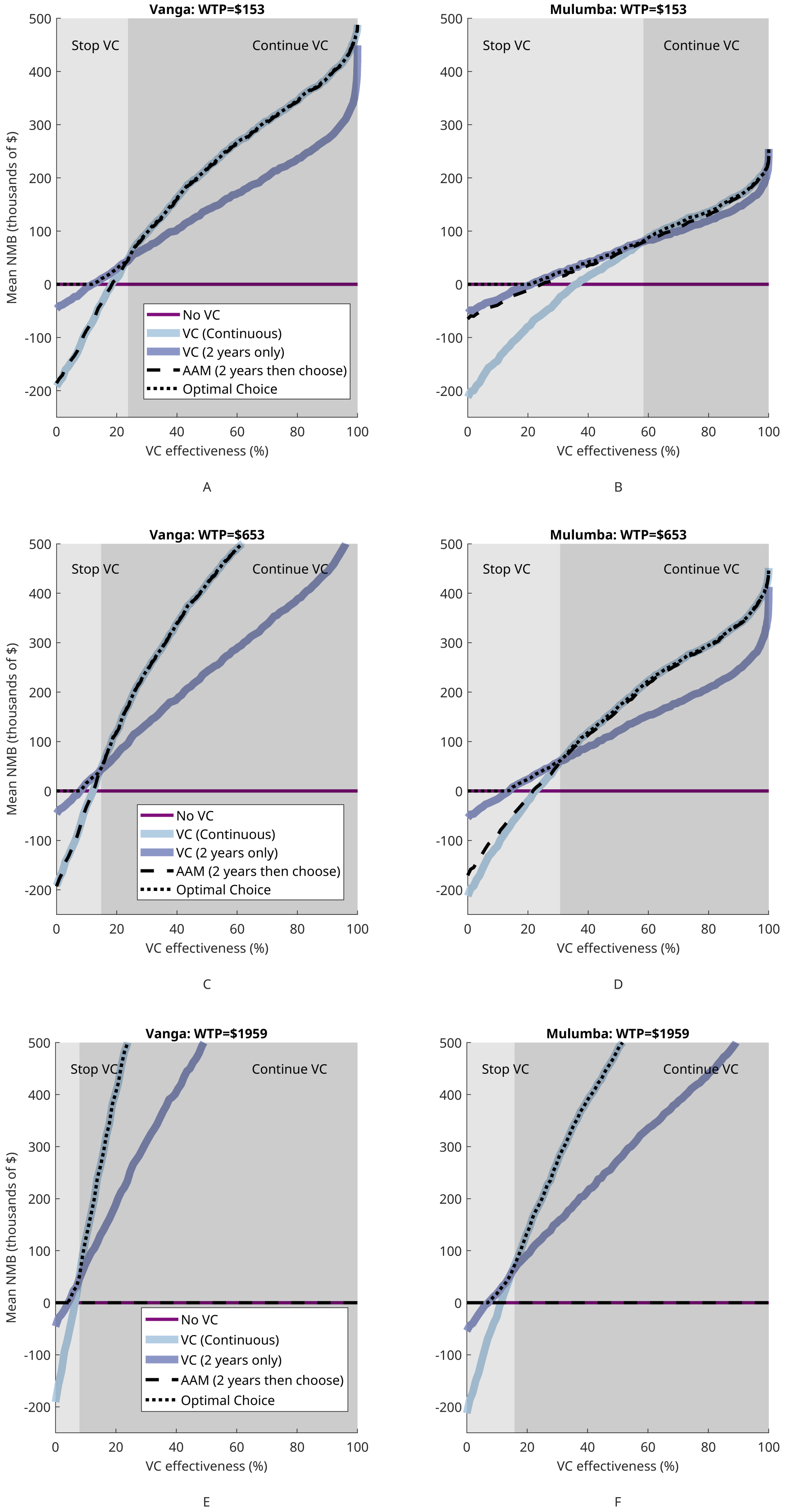


**Fig K.** NMB for a range of different WTP values

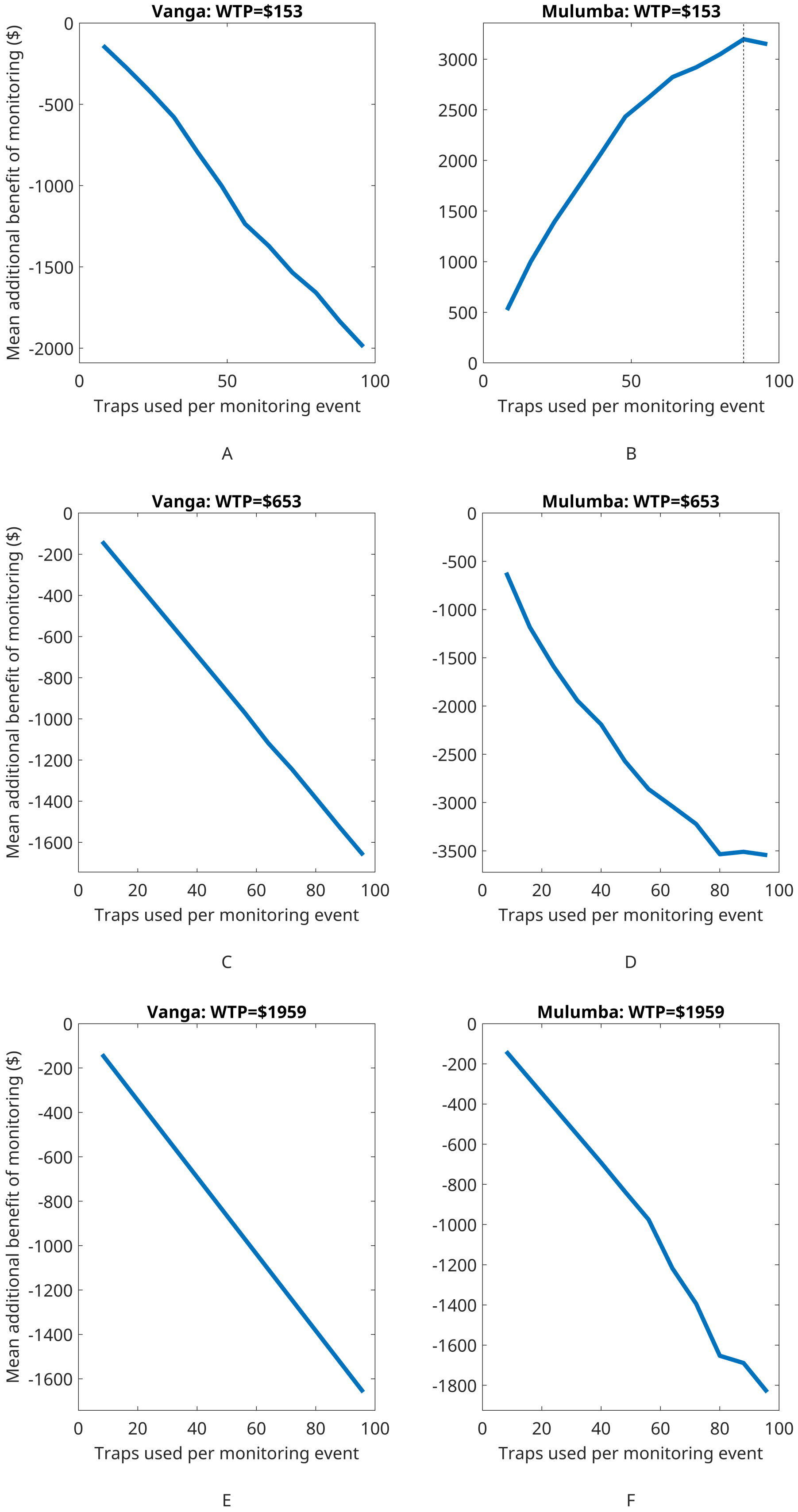


**Fig L.** Benefit of undertaking monitoring for a range of different WTP values

##### S1.6.3 Lower levels of overdispersion

The large amount of overdispersion in the data leads to a lot of uncertainty around decision-making, as evident in the plots that show the probability of choosing to continue VC. To demonstrate the effect that this overdispersion has on our results, we have rerun the same analysis again, but this time using an overdispersion prior that is approximately four times lower:

$$\text{Main Text: Overdispersion} \sim \Gamma(124.7245, \tfrac{1}{0.046115})$$

$$\text{Lower Level: Overdispersion} \sim \Gamma(\tfrac{124.7245}{4}, \tfrac{1}{0.046115})$$

With this new prior on overdispersion, we can again calculate the probability of choosing to continue VC, and how this varies with the true reduction in tsetse, as well as recalculating the benefit of monitoring. In Figure M, we can see that lower levels of overdispersion lead to decision-making that is closer to the optimal choice, and in Figure N we can see how this affects the benefit of monitoring. In Vanga, there is little effect, as whilst decision-making is more likely to determine when VC effectiveness is low, the decision to continue VC is still very dominant. In Mulumba however, we can see that a lower level of overdispersion drastically shifts the benefit curve. Not only does the benefit of monitoring increase due to the better decision-making, but the optimal number of traps is now lower, as we can obtain more confident posterior distributions with less data.

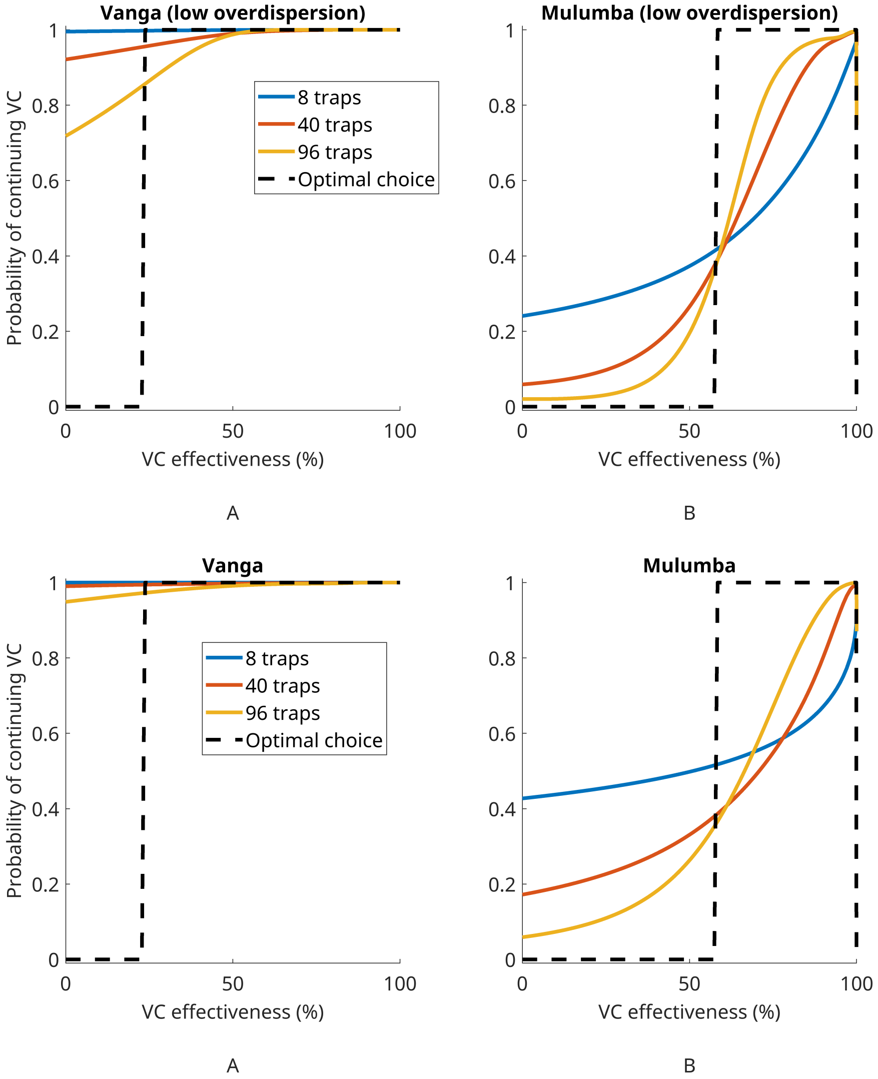


**Fig M.** The probability of choosing to continue VC versus the true reduction in the tsetse population. We show this for a range of different numbers of traps as well as showing the optimal choice. The top row is what we expect with lower overdispersion, whilst the bottom row is what we obtained from our standard overdispersion prior used in the main text.

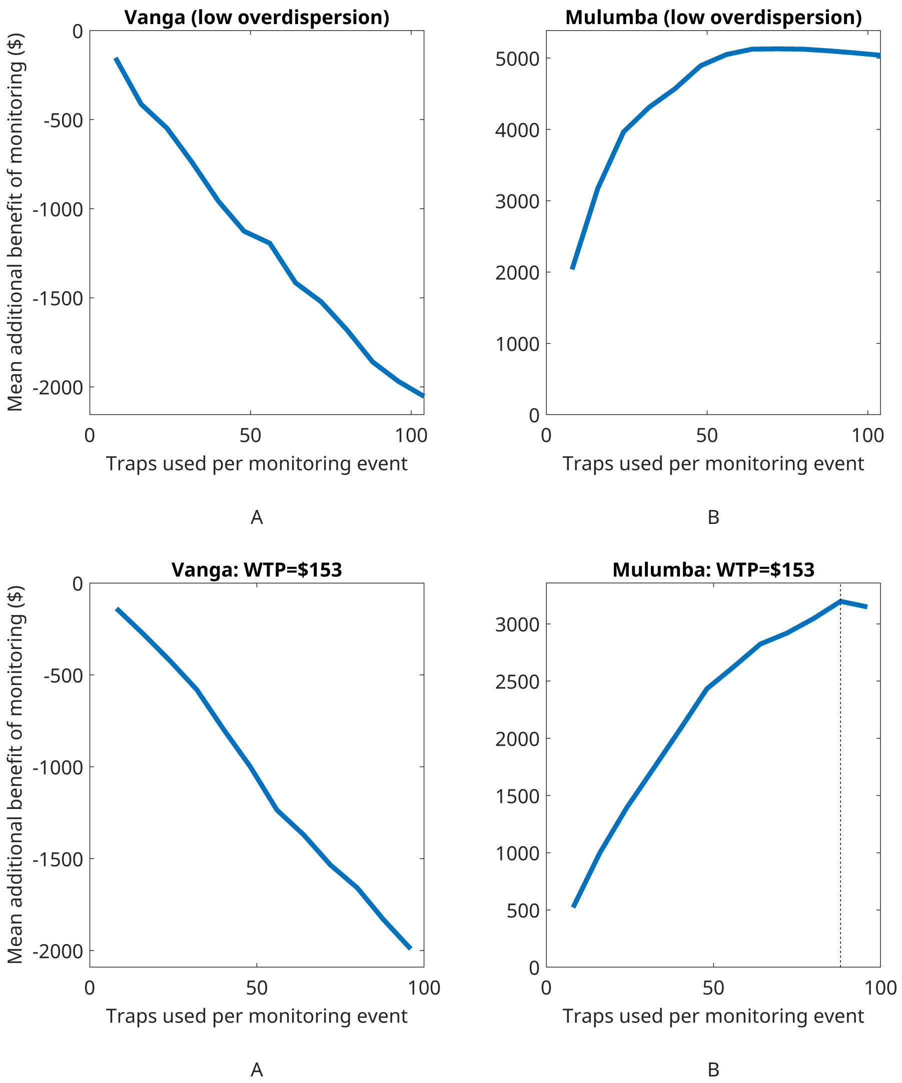


**Fig N.** The probability of choosing to continue vector control versus the true reduction in the tsetse population. We show this for a range of different numbers of traps as well as showing the optimal choice. The top row is what we expect with lower overdispersion, whilst the bottom row is what we obtained from our standard overdispersion prior used in the main text. Willingness to pay is $153 in both.

#### References


1. Antillon M, Huang CI, Sutherland SA, Crump RE, Brown PE, Bessell PR, et al. Cost-effectiveness of end-game strategies against sleeping sickness across the Democratic Republic of Congo. medRxiv. 2024;doi:10.1101/2024.03.29.24305066.

2. Sunnucks R, Davis EL, Rock KS. Methods for Reproducible Comparison of Strategies in Stochastic Modelling. medRxiv. 2025;doi:10.1101/2025.10.09.25337145.

3. Rock KS, Torr SJ, Lumbala C, Keeling MJ. Quantitative evaluation of the strategy to eliminate human African trypanosomiasis in the Democratic Republic of Congo. Parasites & vectors. 2015;8(1):532.

4. Crump R, Aliee M, Sutherland S, Huang C, Crowley E, Spencer S, et al. Modelling timelines to elimination of sleeping sickness in the Democratic Republic of Congo, accounting for possible cryptic human and animal transmission. Parasit Vectors. 2024;doi:https://doi.org/10.1186/s13071-024-06404-4.

5. Crump RE, Huang CI, Knock ES, Spencer SEF, Brown PE, Mwamba Miaka E, et al. Quantifying epidemiological drivers of gambiense human African Trypanosomiasis across the Democratic Republic of Congo. PLOS Computational Biology. 2021;17(1):1–23. doi:10.1371/journal.pcbi.1008532.

6. Tirados I, Hope A, Selby R, Mpembele F, Miaka EM, Boelaert M, et al. Impact of tiny targets on Glossina fuscipes quanzensis, the primary vector of human African trypanosomiasis in the Democratic Republic of the Congo. PLOS Neglected Tropical Diseases. 2020;14(10):1–20. doi:https://doi.org/10.1371/journal.pntd.0008270.

7. Davis CN, Crump RE, Sutherland SA, Spencer SEF, Corbella A, Chansy S, et al. Comparison of stochastic and deterministic models for gambiense sleeping sickness at different spatial scales: A health area analysis in the DRC. PLOS Computational Biology. 2024;20(4):1–25. doi:10.1371/journal.pcbi.1011993.

8. Rock KS, Torr SJ, Lumbala C, Keeling MJ. Predicting the impact of intervention strategies for sleeping sickness in two high-endemicity health zones of the Democratic Republic of Congo. PLoS neglected tropical diseases. 2017;11(1):e0005162.

9. Antillon M, Huang CI, Crump RE, Brown PE, Snijders R, Miaka EM, et al. Cost-effectiveness of sleeping sickness elimination campaigns in five settings of the Democratic Republic of Congo. Nat Commun. 2022;doi:https://doi.org/10.1038/s41467-022-28598-w.

10. Woods B, Revill P, Sculpher M, Claxton K. Country-Level Cost-Effectiveness Thresholds: Initial Estimates and the Need for Further Research. Value in Health. 2016;19(8):929–935. doi:https://doi.org/10.1016/j.jval.2016.02.017.

11. in2013dollars com. Inflation Calculator;. Available from: `https://www.in2013dollars.com/us/inflation/2016?endYear=2022&amount=1`.

12. Bank W. Congo [DRC] Data Commons;. Available from: `https://datacommons.org/place/country/COD?utm_medium=explore&mprop=amount&popt=EconomicActivity&cpv=activitySource,GrossDomesticProduction&hl=en#`.

13. Snijders R, Shaw APM, Selby R, Tirados I, Bessell PR, Fukinsia A, et al. The cost of sleeping sickness vector control in the Democratic Republic of the Congo. medRxiv. 2024;doi:10.1101/2024.02.02.24302172.

14. Kaba D, Djohan V, Berte D, TA BTD, Selby R, Kouadio KADM, et al. Use of vector control to protect people from sleeping sickness in the focus of Bonon (Côte d'Ivoire). PLOS Neglected Tropical Diseases. 2021;15(6):1–18. doi:10.1371/journal.pntd.0009404.
